# An epidemiological study of season of birth, mental health, and neuroimaging in the UK Biobank

**DOI:** 10.1101/2023.08.09.23293866

**Authors:** Maria Viejo-Romero, Heather C. Whalley, Xueyi Shen, Aleks Stolicyn, Laura de Nooij, Daniel J. Smith, David M. Howard

## Abstract

Environmental exposures during the perinatal period are known to have a long-term effect on adult physical and mental health. One such influential environmental exposure is the time of year of birth which affects the amount of daylight, nutrients, and viral load that an individual is exposed to in the key developmental period. Here we investigate associations between season of birth (seasonality), four mental health traits (*n*=135,541) and multi-modal neuroimaging measures (*n*=33,815) within the UK Biobank. Summer births were associated with probable recurrent Major Depressive Disorder (β=0.024, *p_corr_=*0.048), greater mean cortical thickness in temporal and occipital lobes and in the middle temporal, fusiform, superior temporal, and lingual gyri regions (β range=0.013 - 0.020, *p_corr_<*0.05). Winter births were associated with greater white matter integrity globally, in the association fibers, thalamic radiations, and six individual tracts (β range=-0.010 to -0.021, *p_corr_<*0.05). Results of sensitivity analyses that adjusted for birth weight were similar, with additional associations found between winter birth and frontal, occipital and cingulate lobe surface areas, as well as fractional anisotropy in the forceps minor. Sensitivity analyses also revealed an additional association between summer birth and greater cingulate thickness. Overall, results suggest that seasonality affects brain structure in later life and may have a role in lifetime recurrent Major Depressive Disorder. The small effect sizes observed here warrant further research to validate the results in the context of different latitudes and co-examine genetic and epigenetic measures to potentially reveal informative biological pathways.

## Introduction

Season of birth has long been hypothesised to have enduring effects on human health [1]. The relatively recent “Foetal Origins of Adult Disease” hypothesis proposes that intra-uterine exposures influence later adult health, including mental health outcomes [2, 3]. Seasonality of birth has been associated with multiple diseases [4], including psychiatric [5-7], neurodevelopmental [8], cardiovascular [9], inflammatory [10], and genetic [11]. The mechanisms by which seasonality may affect risk to these disorders are hypothesised to include interactions with photoperiod and sunlight [12], nutrition [13], risk of pre-term birth and early life infection [14], and maternal vitamin D deficiency [15]. These interactions could also be mediated by genetic [16], epigenetic [17] or environmental changes [5] i*n utero* or perinatally.

Perinatal photoperiod directly affects the physiology, brain morphology and behaviour of many animals via transplacental signalling to melatonin receptors in the developing medio-basal hypothalamus [18, 19]. Many of these season-of-birth effects endure into adulthood, including altered circadian timing [20], changes in affective behaviour, hippocampal volume [21] and changes to serotonergic and dopaminergic systems in the brain [22]. The wider effects of season of birth on human brain structure have been investigated using magnetic resonance imaging with winter births associated with increased grey matter volume of the superior temporal gyrus in a study of over 550 individuals [23]. However, in a larger study of 13,000 Rotterdam Study participants there was no effect of season of birth on any imaging derived measures [24].

Differences in photoperiod exposure during the perinatal and postweaning time windows have been associated with enduring changes in anxiety and depressive-like behaviours in developmentally stable adulthood periods within animal models [25, 26], with shorter photoperiods associated with increased presence and severity of these behaviours. Differences in brain morphology have been associated with mental health disorders including depression [27-29], externalizing behaviour [30] and schizophrenia [31, 32]. The associations between brain morphology and mental health thus warrant an investigation of associations of seasonality with both brain morphology and mental health.

Birth seasonality likely plays a multifaceted role in the aetiology of adult mental health, either alongside or independently of seasonality-induced brain morphology changes. However, to date, the potential relationship between seasonality of birth, mental health traits and imaging measures has not been examined at scale. This study, therefore, aims to individually explore associations between seasonality of birth and (a) mental health disorders, and (b) brain imaging measures within the UK Biobank.

## Materials & Methods

### UK Biobank

The UK Biobank (UKB) is a well-characterised community cohort of over 500,000 participants aged 37-73 years at recruitment (2006-2010) [33]. All participants were invited to an initial assessment in which baseline data were collected, including month of birth, birth weight, and birth location. Mental health information was collected for all individuals at baseline using a Touchscreen Questionnaire, and for 157,348 participants between 2016 and 2017 using a Mental Health Questionnaire [34] (MHQ). A subset of the cohort (*n*=42,709) was invited to the first imaging visit in 2014 where brain scans, including T1-weighted (T1) and diffusion tensor imaging (DTI), were obtained. Data were accessed under the UKB Application Number 4844. UKB has approval from the NHS National Research Ethics Service as a research tissue bank (References 16/NW/0274 and 11/NW/0382).

### Seasonality

Seasonality of birth was examined as a quantitative phenotypic trait (*y*) capturing the month of birth (UKB data-field 52) following the approach of Howard et al. (35). The birth month of each participant (*i*) was transformed via a cos function, with the lowest phenotypic score (-1) corresponding to those born in December and the highest phenotypic score corresponding to those born in June (+1), as below:

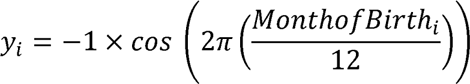

### Mental health traits

Four mental health trait phenotypes were obtained from responses to the Touchscreen Questionnaire and the “Thoughts and Feelings” section of the MHQ. Where participants had completed both questionnaires (*n*=85,266), the MHQ responses were used to generate phenotypes due to the greater specificity of this questionnaire (see S2.2.1A and S2.2.1B Methods for UKB data-fields used). Participants who had answered “Prefer not to answer” or “I don’t know” to any questions or with insufficient symptom data (i.e., only one reported symptom) were removed at this stage. The remaining participants (*n*=170,982) were categorised into four mental health-related phenotypes: probable recurrent Major Depressive Disorder (P-RMDD) (*n*=39,528 cases), probable single episode Major Depressive Disorder (P-SEMDD) (*n*=16,430 cases), probable Hypomania (*n*=9,104 cases) and probable Mania (*n*=2,569 cases), plus a control group (*n*=103,351) as previously validated by Smith et al. (36) (see S2.2.1C and S2.2.1D Methods for grouping specification). At this point, participants who were of non-white ethnicity (*n*=9,581), did not report a UK or Republic of Ireland birthplace or had unrecoverable geographical birthplace co-ordinate data (*n*=9,897) (See S2.1 Methods for geographical data processing), had missing Townsend Deprivation Index values (*n*=189), or had chosen to withdraw from further studies (*n*=8) were excluded. Furthermore, participants who reported other neuropsychiatric or sleep-related conditions (*n*=28), brain cancer diagnoses (*n*=2), or reported having done shift work (*n*=13,241) were also excluded. This resulted in a total of 32,946 exclusions with 138,036 participants remaining (see S2.3.1A Methods for exclusion details).

Any overlaps between the groupings were removed to create distinct phenotypes that did not share a probable diagnosis: P-RMDD (*n*=31,652), P-SEMDD (*n*=13,529), probable Unipolar Mania (P-UM) (*n*=1,202) and probable Bipolar Depression (P-BD) (*n*=5,140) (see S2.2.1D Methods for overlaps permitted). Both the P-RMDD and the P-SEMDD groups exclude participants within the probable Mania or probable Hypomania groups. P-UM excludes participants within either depression group, whereas the P-BD grouping allows for participants also within either depression grouping and follows the definitions used in Sangha et al. (37). Participants who completed the MHQ lead questions for mania (UKB data-fields 20501 and 20502) and depression (UKB data-fields 20446, 20441), and/or the Touchscreen Questionnaire lead questions for mania (UKB data-fields 4642, 4653) and depression (UKB data-fields 4598, 4631) but had not been classified into one of the four phenotypes, were defined as controls (*n*=84,018). The total sample size was therefore 135,541 (see S2.3.1B Methods for group sample sizes). Participant demographics for the mental health phenotypes are provided in Table 1.

**Table 1.** Participant demographic details for mental health trait analysis. SD = Standard deviation. Birth location was obtained using k-means clustering (see Statistical models section for full details)

|  | Control<br>(N=84018) | Probable Major<br>Depressive<br>Disorder<br>(n=13529) | Probable Single<br>Episode<br>Depression<br>(n=31652) | Probable<br>Unipolar<br>Mania<br>(n=1202) | Probable<br>Bipolar<br>Depression<br>(n=5140) | Total<br>(n=135541) |
| --- | --- | --- | --- | --- | --- | --- |
| **Sex** |  |  |  |  |  |  |
| Female | 39245 (46.7%) | 9306 (68.8%) | 21329 (67.4%) | 460 (38.3%) | 3039 (59.1%) | 73379 (54.1%) |
| Male | 44773 (53.3%) | 4223 (31.2%) | 10323 (32.6%) | 742 (61.7%) | 2101 (40.9%) | 62162 (45.9%) |
| **Age (years)** |  |  |  |  |  |  |
| Mean (SD) | 62.8 (8.32) | 61.5 (8.09) | 60.3 (8.28) | 61.2 (8.42) | 59.8 (7.81) | 62.0 (8.35) |
| **Ethnicity** |  |  |  |  |  |  |
| White | 62 (0.1%) | 7 (0.1%) | 29 (0.1%) | 2 (0.2%) | 8 (0.2%) | 108 (0.1%) |
| British | 81900 (97.5%) | 13129 (97.0%) | 30522 (96.4%) | 1152 (95.8%) | 4908 (95.5%) | 131611 (97.1%) |
| Irish | 1222 (1.5%) | 225 (1.7%) | 591 (1.9%) | 23 (1.9%) | 131 (2.5%) | 2192 (1.6%) |
| Any other white<br>background | 834 (1.0%) | 168 (1.2%) | 510 (1.6%) | 25 (2.1%) | 93 (1.8%) | 1630 (1.2%) |
| **Month of Birth** |  |  |  |  |  |  |
| January | 6894 (8.2%) | 1140 (8.4%) | 2666 (8.4%) | 108 (9.0%) | 443 (8.6%) | 11251 (8.3%) |
| February | 6686 (8.0%) | 1079 (8.0%) | 2474 (7.8%) | 93 (7.7%) | 380 (7.4%) | 10712 (7.9%) |
| March | 7655 (9.1%) | 1241 (9.2%) | 2928 (9.3%) | 109 (9.1%) | 449 (8.7%) | 12382 (9.1%) |
| April | 7264 (8.6%) | 1172 (8.7%) | 2794 (8.8%) | 123 (10.2%) | 429 (8.3%) | 11782 (8.7%) |
| May | 7572 (9.0%) | 1209 (8.9%) | 2915 (9.2%) | 99 (8.2%) | 457 (8.9%) | 12252 (9.0%) |
| June | 6952 (8.3%) | 1135 (8.4%) | 2787 (8.8%) | 111 (9.2%) | 447 (8.7%) | 11432 (8.4%) |
| July | 7161 (8.5%) | 1143 (8.4%) | 2696 (8.5%) | 113 (9.4%) | 434 (8.4%) | 11547 (8.5%) |
| August | 6917 (8.2%) | 1164 (8.6%) | 2534 (8.0%) | 96 (8.0%) | 427 (8.3%) | 11138 (8.2%) |
| September | 6920 (8.2%) | 1128 (8.3%) | 2536 (8.0%) | 92 (7.7%) | 464 (9.0%) | 11140 (8.2%) |
| October | 6741 (8.0%) | 1078 (8.0%) | 2465 (7.8%) | 87 (7.2%) | 410 (8.0%) | 10781 (8.0%) |
| November | 6483 (7.7%) | 983 (7.3%) | 2373 (7.5%) | 91 (7.6%) | 385 (7.5%) | 10315 (7.6%) |
| December | 6773 (8.1%) | 1057 (7.8%) | 2484 (7.8%) | 80 (6.7%) | 415 (8.1%) | 10809 (8.0%) |
| **Birth Location** |  |  |  |  |  |  |
| Cluster 1 | 39860 (47.4%) | 6404 (47.3%) | 15039 (47.5%) | 556 (46.3%) | 2410 (46.9%) | 64269 (47.4%) |
| Cluster 2 | 8600 (10.2%) | 1438 (10.6%) | 3225 (10.2%) | 110 (9.2%) | 548 (10.7%) | 13921 (10.3%) |
| Cluster 3 | 15088 (18.0%) | 2459 (18.2%) | 5477 (17.3%) | 226 (18.8%) | 910 (17.7%) | 24160 (17.8%) |
| Cluster 4 | 20470 (24.4%) | 3228 (23.9%) | 7911 (25.0%) | 310 (25.8%) | 1272 (24.7%) | 33191 (24.5%) |

### Brain imaging measures

T1-weighted and DTI brain scans were obtained for 42,709 participants on their first imaging visit [38], with complete brain imaging data available for 37,048 participants. Participants with outlier values in global measures of cortical surface area (*n*=112), mean cortical thickness (*n*=217), cortical volume (*n*=117), subcortical volume (*n*=90), mean diffusivity (MD) (*n*=105), or fractional anisotropy (FA) (*n*=232) were excluded. Global measures were derived by conducting principal component analyses (PCA) on data from the entire sample [39], and measure outliers were defined as values ±3 standard deviations from the sample mean for that measure. Participants of non-white ethnicity *(n=*1,168), non-UK/Republic of Ireland, non-specified (*n*=1,874) or unrecoverable geographical (*n*=581) birthplace, with a missing Townsend Deprivation Index value (*n*=26), or who wished to withdraw from future studies (*n*=4) were also excluded. No participants had to be removed due to head motion in the scanner. A total of 32,815 participants were included after 3,651 exclusions. Demographics of participants included in the analyses of brain imaging measures are provided in Table 2. An additional sensitivity analysis was conducted using only individuals with birth weight data (*n*=21,182).

**Table 2.** Participant demographic details for neuroimaging analysis. SD = Standard deviation. Birth location was obtained using k-means clustering (see Statistical models section for full details)

|  | Total<br>(n=32815) |
| --- | --- |
| **Sex** |  |
| Female | 17519 (53.4%) |
| Male | 15296 (46.6%) |
| **Age (years)** |  |
| Mean (SD) | 63.7 (7.43) |
| **Ethnicity** |  |
| White | 16 (0.0%) |
| British | 31812 (96.9%) |
| Irish | 578 (1.8%) |
| Any other white background | 409 (1.2%) |
| **Birth Weight (kg)** |  |
| Mean (SD) | 3.36 (0.614) |
| Median [Min, Max] | 3.37 [0.740, 6.78] |
| **Month of Birth** |  |
| January | 2731 (8.3%) |
| February | 2669 (8.1%) |
| March | 3018 (9.2%) |
| April | 2864 (8.7%) |
| May | 2873 (8.8%) |
| June | 2754 (8.4%) |
| July | 2802 (8.5%) |
| August | 2709 (8.3%) |
| September | 2673 (8.1%) |
| October | 2640 (8.0%) |
| November | 2445 (7.5%) |
| December | 2636 (8.0%) |
| **Birth Location** |  |
| Cluster 1 | 6046 (18.4%) |
| Cluster 2 | 11871 (36.2%) |
| Cluster 3 | 2644 (8.1%) |
| Cluster 4 | 12254 (37.3%) |
| **Scanner Site** |  |
| Cheadle | 20111 (61.3%) |
| Reading | 3954 (12.0%) |
| Newcastle | 8750 (26.7%) |

### Brain morphology measures

Brain morphometric measures were obtained by the UK Biobank with FreeSurfer 6.0 toolkit [40-42] and included volumes of seven subcortical structures for each hemisphere, as well as volume, thickness and surface area measures of 31 cortical regions for each hemisphere, based on the Desikan-Killiany-Tourville atlas [43] (n=32,815, see S1.1 Methods for full pre-analysis QC). Similar measures were analysed in Harris et al. (39) (UKB category 192) and Shen et al. (29). Global and lobar cortical thickness, surface area and cortical volume measures were derived manually for each hemisphere (n=32,815). Lobar measures were obtained for the frontal, parietal, temporal, occipital and cingulate lobes (see S2.2.2A Methods for group composition). All brain morphometric measures were normalised.

### White matter microstructure measures

White matter (WM) microstructure measures consisted of FA and MD values for 12 bilateral tracts and 3 unilateral tracts derived by the UK Biobank with the FSL probabilistic tractography toolkit (See S1.1 Methods) [38, 44]. Additional fiber-related FA and MD measures were derived as the scores on the first unrotated principal components from PCA, which combined bi-hemispheric (left and right) and unilateral measures from all relevant individual fiber tracts. The three whole-brain fiber bundles derived were the association fibers, projection fibers and thalamic radiations (see Methods S2.2.2A for fiber definitions). Global FA and MD measures were derived as the scores on the first unrotated principal components from PCA analyses which combined all bi-hemispheric and unilateral tract measures. Proportions of variance explained by the first principal components are provided in Methods S2.2.2B. Thirty-eight WM integrity measures were analysed in total (19 FA and 19 MD) (*n*=32,815) and all WM microstructure measures were normalised.

## Statistical models

### Mental health traits

To investigate associations between the four mental health phenotypes and seasonality, logistic binomial regression analyses were performed, covarying for sex, age, age ^2^, Townsend Deprivation Index, assessment centre attended and place of birth location. Place of birth locations were derived with k-means clustering of participant birth north / east co-ordinates from the Ordnance Survey data (UKB variables 129 and 130), performed using “kclust” function in the R “stats” package. Twenty clustering iterations were run to identify four birth location clusters (see S2.1.1 Methods and S1-S3 Fig), and each participant was assigned to a cluster to define place of birth. Sex, assessment centre and place of birth were coded as categorical variables. A Bonferroni multiple analysis correction was applied over the four phenotypes examined (P < 0.0125 (α = 0.05 / 4)). Effect sizes were standardised throughout.

### Brain imaging measures

Linear regression models were applied to assess associations between seasonality and all unilateral, fiber-related or global brain measures. Mixed-effects models were applied to assess associations between seasonality and all bilateral brain measures (“nlme” package in R version 3.2.3). Sex, age, age^2^, Townsend Deprivation Index, assessment centre, four UKB head position covariates (X, Y, Z and table position) and place of birth cluster index (see S4-S6 Fig) were included as covariates in all analyses. Sex, assessment centre and place of birth were coded as categorical variable Hemisphere was controlled for as a random effect in the mixed-effect models for all bi-hemispheric measures (see S2.1.2 Methods). Standardised intra-cranial volume was also covaried for in all analyses of brain morphometric measures.

False Discovery Rate (FDR) correction was applied separately across individual and regional brain morphology and white matter microstructure measures per modality (Methods S2.4.1). Global measures were not corrected. A P-value threshold for significance was set to 0.05 and effect sizes were standardised throughout. All statistical analyses were performed using R (version 3.2.3). A sensitivity analysis was conducted to reassesses the brain imaging measures for an association with seasonality after fitting birth weight as an additional fixed effect covariate.

## Results

### Seasonality associations with mental health traits

Probable recurrent Major Depressive Disorder was associated with seasonality, with a higher prevalence observed in summer births (β= 0.024, *p_corr_=* 0.048). No other mental health traits were associated with seasonality (Table 3). Effect sizes are reported as log-transformed odd ratios.

**Table 3.** Mental health traits associated with seasonality. p-uncorr = p-uncorrected value; p-corr = Bonferroni p-corrected value; S.E = standard error.

| Mental Health Trait | Effect Size($\beta$)<br>/ Log(OR) | S.E. | p-uncorr | p-corr |
| --- | --- | --- | --- | --- |
| Probable recurrent Major Depressive Disorder | 0.024 | 0.010 | 0.012 | 0.048 |
| Probable single Episode Major Depressive Disorder | 0.016 | 0.013 | 0.243 | 0.973 |
| Probable Unipolar Mania | 0.061 | 0.041 | 0.139 | 0.554 |
| Probable Bipolar Depression | 0.007 | 0.021 | 0.754 | 1.000 |

### Seasonality associations with brain imaging measures

Greater mean temporal lobe thickness (β= 0.011, *p_corr_=* 0.017) and greater mean occipital lobe thickness (β= 0.010, *p_corr_=* 0.025) were associated with summer births. Summer births were also associated with four individual regional cortical thickness measures: middle temporal gyrus (β= 0.018, *p_corr_=* 0.027), fusiform gyrus (β= 0.018, *p_corr_=* 0.031), superior temporal gyrus (β= 0.016, *p_corr_=* 0.033), and lingual gyrus (β= 0.016, *p_corr_=* 0.049) (Fig 1).

**Fig 1.**
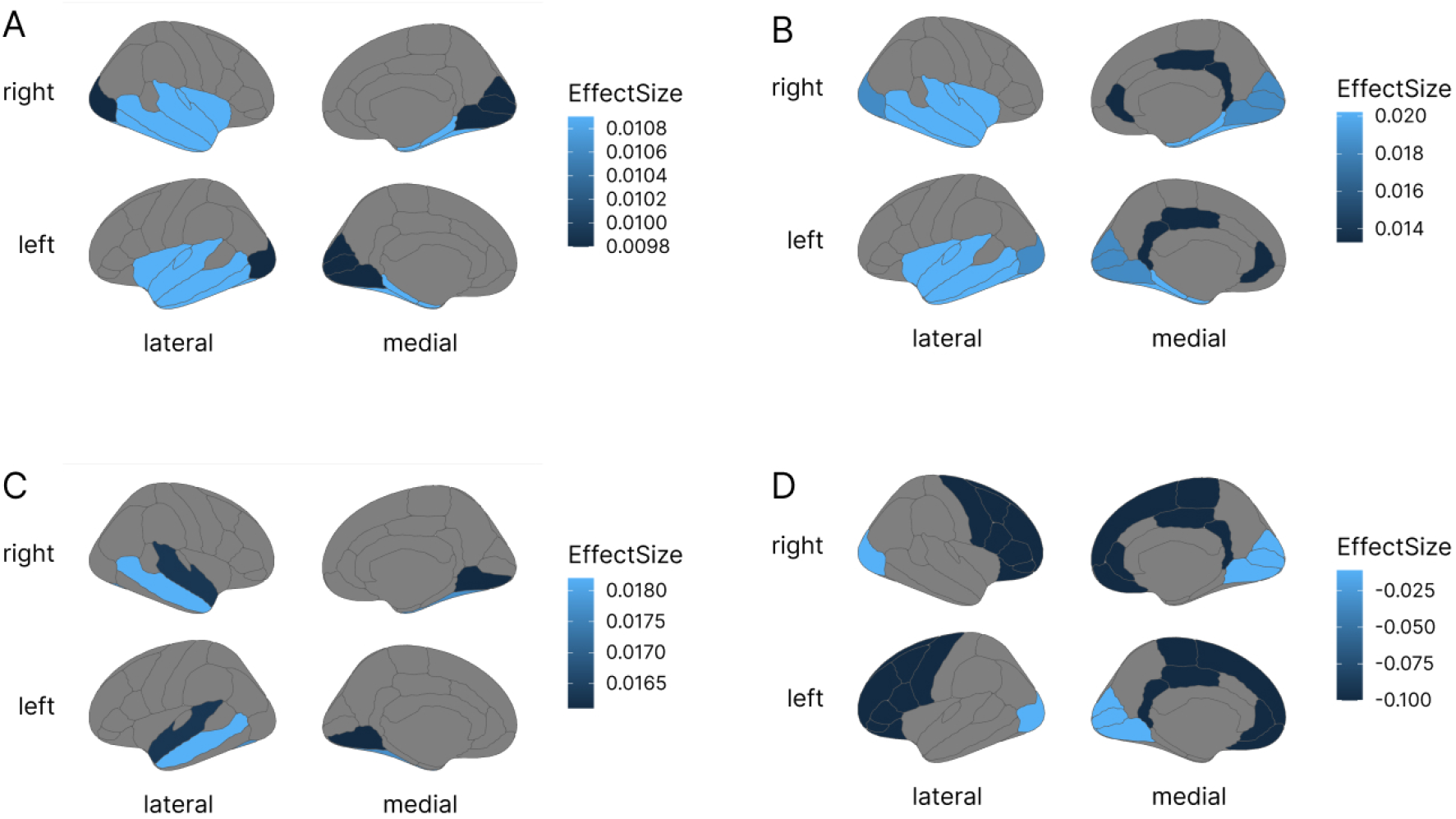
Standardised effect sizes of brain morphology measures associated with seasonality mapped onto the Desikan-Killiany-Tourville atlas for (A) mean cortical thickness of the temporal and occipital lobes in the main analyses and in the sensitivity analyses for (B) mean cortical thickness of the middle temporal, fusiform, superior temporal and lingual gyri, (C) mean cortical thickness of the temporal, occipital and cingulate lobes, and (D) cortical surface area of the frontal, occipital and cingulate lobes. Darker colour indicates associations with winter births, lighter colour indicates associations with summer births.

Winter births were associated with higher global FA (β= -0.013, *p*=0.001), higher FA in the association and thalamic fibers (respectively β= -0.016, *p_corr_=* 2.63 × 10^-5^ and β= -0.011, *p_corr_=* 0.004), and higher FA in six of 15 individual WM tracts (effect sizes ranging from β= -0.009 to β= -0.016) (Fig 2, S3.1 Results). No MD measures were associated with seasonality.

**Fig 2.**
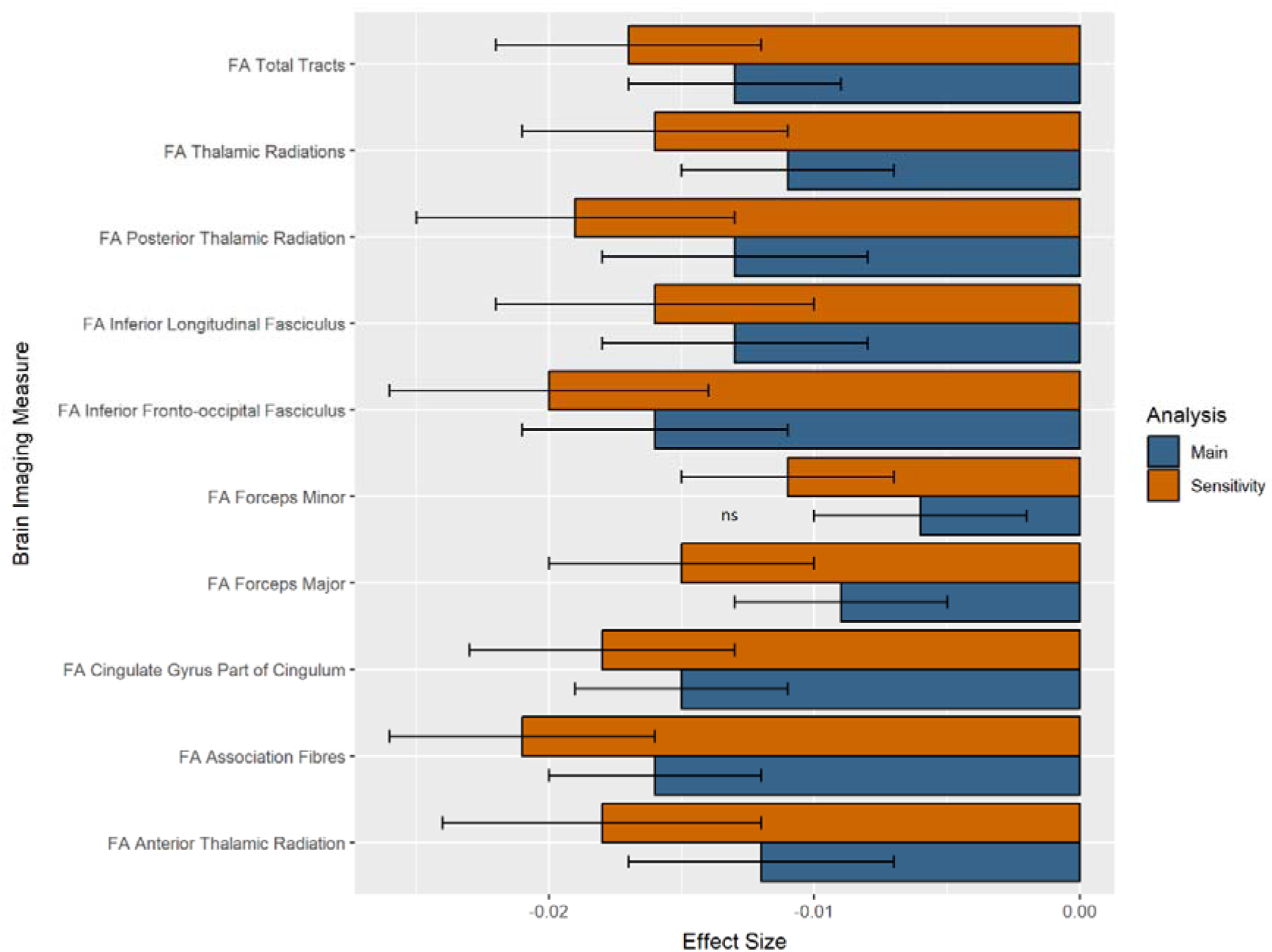
Standardised effect sizes for white matter microstructure neuroimaging measures associated with seasonality in the main and sensitivity analyses. All shown measures were associated with seasonality (p_corr_<0.05), except for FA Forceps Minor in the main analysis (ns = non-significant).

When birth weight was additionally covaried for in the sensitivity analyses, four additional cortical morphometric measures were identified as significantly associated with seasonality: frontal lobe surface area (β= -0.010, *p_corr_=* 0.025), occipital lobe surface area (β= -0.011, *p_corr_=* 0.025), cingulate surface area (β= -0.010, *p_corr_=* 0.026), and cingulate cortical thickness (β= 0.013, *p_corr_=* 0.007) (Fig 1). Four previously identified associations remained significant with greater observed effect sizes (range β= 0.013 to β= 0.020) (see S3.2 Results) With regard to WM measures, an additional association of FA in the forceps minor with winter births was identified when correcting for birth weight (β= -0.011, *p_corr_=* 0.029), while all previous associations remained significant with larger effect sizes (see Fig 2, S3.2.5-S3.2.7 Results for further details).

## Discussion

This study investigated associations between seasonality and mental health traits and neuroimaging measures in a large-scale cross-sectional dataset. Seasonality was associated with probable recurrent Major Depressive Disorder, as well as with a range of brain morphology and white matter microstructure brain imaging measures. Summer births were associated with a higher prevalence of probable recurrent Major Depressive Disorder, as well as greater mean thickness in the temporal, occipital and cingulate lobes, and the middle temporal, fusiform, superior temporal, and lingual gyri. Winter births were associated with more constrained water molecule diffusion and thus higher white matter integrity globally, in two white matter fiber tract bundles and in seven individual white matter tracts, as well as greater area measures in the frontal, occipital and cingulate areas. These results lend additional support to existing evidence of season of birth effects in adult health.

In this study P-RMDD was associated with summer births, aligning with northern hemisphere spring associations with MDD risk in English outpatients (*n*=16,726) [6], severity in a small MDD case-control study (45 cases and 90 controls) [45] and earlier disease onset in 855 Korean MDD patients [46]. However, there exist idiosyncrasies in the seasonality phenotypes derived in these studies, which range from monthly, to two-and four-season seasonality categories per participant, making for inexact comparison. A hemispheric six-month shift has also been observed between seasonality and depression symptoms, with higher scores associating with spring births (March-May) in the Northern hemisphere and autumn births (September-November) in the Southern hemisphere, in a small young adult and adolescent cohort [47]. Although an excess of August births in MDD patients has also been observed in the Southern hemisphere within a Brazilian retrospective study of MDD cases and controls (*n*=98,457) [48]. However, no associations between month of birth and depression symptoms were found in a similarly aged cohort in a recent Europe wide study (*n*=72,370) [49]. For P-RMDD, therefore, our findings provide a seasonality association within a UK population sample adding another datapoint to the ambiguous literature.

Unlike P-RMDD, single episode MDD was not associated with seasonality in this study. The more than two-fold reduction in sample size for P-SEMDD (*n=*13,529) may have limited statistical power to detect associations. An unshared aetiology may also be a factor, with more severe neurophysiological observations made in clinically comparable recurrent MDD cases versus single episode MDD cases [50], and an earlier age of onset and familial risk for recurrent MDD [51, 52]. Gene ⍰ environment effects [53] and genetic variants [54, 55] associated with recurrent MDD support this distinction. Therefore, seasonality effects may vary within depressive subtypes, with a more marked effect on persistent cases in response to seasonal perinatal and natal environments.

Bipolar affective disorder has been associated with January births [6] (OR=1.09, 95% CI= 1.03–1.15, *p* =0.002), and for DSM-III bipolar disorder cases a seasonal pattern has been observed with significant excess births in December and a total of 5.8% seasonal excess births to expected births [56]. However, the current study did not find a seasonal association, possibly due to the inclusion of both P-RMDD and P-SEMDD within P-BD since only the former associated with seasonality. It is also possible that the self-reported phenotype used may not fully capture cases with sufficient fidelity to yield associations. UM, a relatively unexplored BD-subtype [57], had the largest effect size of the mental health traits but was not associated with seasonality potentially due to the small number of cases available. Re-examination of this phenotype in a larger UM sample is therefore warranted.

In line with previous studies, we find seasonality to be associated with a cluster of white matter microstructure measures, with novel findings for brain morphology regional measures. Overall, six regional and four individual brain morphology measures were associated with seasonality, all of which were either in the area or mean thickness category, despite the lack of associations for volumetric measures and seasonality expressed as daylength in similarly powered UKB studies [16]. Thickness of the temporal lobe has been shown to be significantly reduced in schizophrenic patients [58], and the disorder’s own association with winter births [59], may offer an opportunity for further study. Since cortical thickness has been less explored in the field, these associations merit closer study.

The observation of multiple negative associations for DTI FA measures in association with summer births, corroborates previous findings in which sections of the corpus callosum, the internal capsule, the corona radiata, the posterior thalamic radiation and the sagittal striatum were also found to have decreased FA values in summer compared to winter births [60], with the majority of these associations retaining significance in our sensitivity analysis. For summer births, lower FA measures, or less restricted and more isotropic diffusivity, may point to greater tissue disorganisation, itself generally accompanied by reduced axonal myelination, axonal loss, or a higher proportion of crossing fibers. Since global and regional measures yielded the strongest associations, alongside seven individual measures, seasonality may exert a non-localised effect on white matter integrity and thus warrant further analysis to specify possible mechanisms. The overall association of the thalamic radiations bundle and the association fibers bundle, composed of two and three individual tracts with individual associations respectively, provides novel evidence for lower values in summer births.

Birth seasonality effects, therefore, may independently induce penetrable changes in white matter integrity in a subset of tract bundles, an effect discernible through DTI-ascertained measures. Brain structure and connectivity measures are a promising endophenotype for mental, neurological, and physiological illness. The range of neuroimaging associations with seasonality found here offers a starting point for further probing into the mechanistic relationship between them.

This study was limited in geographical scope with the aim of keeping latitudinal and longitudinal variation minimal between subjects. Since season of birth effects have been shown to be greater at higher latitudes [59], possibly mediated by larger annual photoperiod shifts, studies in these regions, or meta-studies encompassing them might provide further insight.

Although our mental health trait phenotypes align well with DSM-5 diagnostic criteria [34], they do not reflect formal diagnoses and therefore may under-or over-extend seasonality associations present under stricter definitions. A symptom-by-symptom study could also elucidate individual patterns in associations, such as MDD sleep aberrances and evidenced seasonality mediated sleep differences [16]. The natural patterns of distribution in birth seasonality [61] with April and May annual birth-rate peaks in UKB [16] should also be accounted for, as should the seasonal pattern in procreation habits observed in psychiatric disorders such as schizophrenia, which may be tied to heritable components [62] of mental health disorders independently. Here, birth seasonality variation was assessed comparing winter births with summer birth. However, this provides those born in spring and autumn with similar phenotypic scores and therefore will not model differences between those born in those periods. Future studies could co-model a range of seasonality phenotypes to contextualise the extent of these potential differences.

Since historically, seasonality effects on psychiatric conditions have been more pronounced in public versus private hospitals [63] and affected by urbanicity and education level, studies that account for these variables could provide a better picture of seasonality associations, although Townsend Deprivation Area may be a sufficient proxy for these measures. Generally, the lag between the time of birth and seasonality-related outcomes in later life makes their association prone to confounders which must be further considered. Overall, the small effect sizes found in this study suggest a limited role for seasonality in adult health, although usage of larger sample sizes and inclusion of more covariates could modify this.

Lastly, although our study supports an association between seasonality and adult health, identification of the mechanisms by which these effects are actualised is beyond the scope of the current study. Longitudinal studies tracking changes in mental health and neuroimaging measures would more precisely quantify within-individual shifts over the lifetime and ease the identification of key developmental periods in which these take place. Since circadian patterns of gene expression have been demonstrated to be weaker in post-mortem human subjects with MDD [64], further studies examining deficiencies in circadian rhythmicity at the transcriptomic level and their associations with mental health traits in the context of month of birth could also be beneficial. A combinatorial approach including genetic and gene expression data would give insight into differential seasonality programming and begin to specify possible biological pathways.

In summary, we demonstrated seasonality of birth associations with adult health as measured by mental health traits and neuroimaging. The small effect sizes of our associations with global, regional, and individual brain imaging measures, as well as probable recurrent Major Depressive Disorder warrants replication in larger and more diverse datasets as well as those offering wider latitudinal ranges. A continuation of the examination of seasonality associations with mental health is also encouraged within higher powered studies or those utilising diagnosed cases.

## Data Availability

All data produced in the present work are contained in the manuscript.
The raw phenotypic data from UK Biobank used in this study are available from: http://www.ukbiobank.ac.uk/.

## Acknowledgements

This research was conducted using the UK Biobank resource, application number 4844. The UK Biobank study was conducted under generic approval from the NHS National Research Ethics Service (approval letter dated June 17, 2011, Ref 11/NW/0382). All participants gave full informed written consent. We would also like to thank the UK Biobank team and all the participants for their collaboration.

## Author Contributions

M.V.R and D.M.H conceptualised the project. Data curation and software code were provided by X.S, A.S, and L.de N. The analysis was performed by M.V.R. Supervision and results interpretation were provided by H.C.W, D.M.H and D.J.S. M.V.R and D.M.H wrote the original manuscript with review and editing provided by all authors.

## Supporting Information

### Supplementary Methods

S1. UKB Data procedures and acquisition
  - S1.1 Brain imaging measures
S2. Data pre-processing
  S2.1 Covariates
    - S2.1.1 Mental health traits
    - S2.1.2 Brain imaging measures
  S2.2 Classification
    - S2.2.1 Mental health traits
      - S2.2.1A Variables utilised to derive probable mania mental health phenotypes
      - S2.2.1B Variables utilised to derive probable depression mental health phenotypes
      - S2.2.1C Criteria for mental health trait phenotype grouping
      - S2.2.1D Overlaps in mental health phenotype groupings
    - S2.2.2 Brain imaging measures
      - S2.2.2A UKB T1 and DTI brain imaging variables used to derive brain imaging measures
      - S2.2.2B Variance explained by the first principal component for DTI PCA
  S2.3 Quality control
    - S2.3.1 Mental health traits
      - S2.3.1A Conditions excluded for mental health traits
      - S2.3.1B Sample size per mental health traits post-exclusions
  S2.4 Multiple testing correction
    - S2.4.1 Multiple testing corrections applied per neuroimaging measure modality

### Supplementary Results

S3. Brain imaging measures supplementary results
  - S3.1 Seasonality associations with brain imaging measures
    - S3.1.1 Global T1 measures
    - S3.1.2 Lobar T1 measures
    - S3.1.3 Individual T1 measures
    - S3.1.4 Subcortical Measures
    - S3.1.5 DTI Global Measures
    - S3.1.6 DTI Grouped Tract Measures
    - S3.1.7 DTI Individual Tract Measures
  - S3.2 Seasonality associations with brain imaging measures covarying for birth weight
    - S3.2.1 Global T1 measures
    - S3.2.2 Lobar T1 measures
    - S3.2.3 Individual T1 measures
    - S3.2.4 Subcortical Measures
    - S3.2.5 DTI Global Measures
    - S3.2.6 DTI Grouped Tract Measures
    - S3.2.7 DTI Individual Tract Measures

### Supplementary Figures

S1 Fig. Visualisation of a maximum of 12 clusters for birth location via k-means clustering analysis on UKB datafields 129 and 130 for participants who had completed the MHQ and/or mania and depression questions in the touchscreen questionnaire.

S2 Fig. Elbow chart of the total within sum of squares by number of clusters (max N=12) for kmeans clustering performed on UKB datafields 129 and 130 for participants who had completed the MHQ and/or mania and depression questions in the touchscreen questionnaire.

S3 Fig. Visualisation of the final four clusters chosen to be the proxy for birth location via k-means clustering analysis on UKB datafields 129 and 130 for participants who had completed the MHQ and/or mania and depression questions in the touchscreen questionnaire.

S4 Fig. Visualisation of a maximum of 12 clusters for birth location via k-means clustering analysis on UKB datafields 129 and 130 for participants who had attended UKB’s imaging assessment.

S5 Fig. Elbow chart of the total within sum of squares by number of clusters (max N=12) for kmeans clustering performed on UKB datafields 129 and 130 for participants who had attended UKB’s imaging assessment.

S6 Fig. Visualisation of the final four clusters chosen to be the proxy for birth location via k-means clustering analysis on UKB datafields 129 and 130 for participants who had attended UKB’s imaging assessment.

