## Supplementary Information for "An epidemiological study of season of birth, mental health, and neuroimaging in the UK Biobank"

**Table of Contents**

Supplementary Methods

S1. UKB Data procedures and acquisition

- S1.1 Brain imaging measures

S2. Data pre-processing

S2.1 Covariates

- S2.1.1 Mental health traits
- S2.1.2 Brain imaging measures

S2.2 Classification

- S2.2.1 Mental health traits
  - S2.2.1A Variables utilised to derive probable mania mental health phenotypes
  - S2.2.1B Variables utilised to derive probable depression mental health phenotypes
  - S2.2.1C Criteria for mental health trait phenotype grouping
  - S2.2.1D Overlaps in mental health phenotype groupings
- S2.2.2 Brain imaging measures
  - S2.2.2A UKB T1 and DTI brain imaging variables used to derive brain imaging measures
  - S2.2.2B Variance explained by the first principal component for DTI PCA

S2.3 Quality control

- S2.3.1 Mental health traits
  - S2.3.1A Conditions excluded for mental health traits
  - S2.3.1B Sample size per mental health traits post-exclusions

S2.4 Multiple testing correction

- S2.4.1 Multiple testing corrections applied per neuroimaging measure modality

Supplementary Results

S3. Brain imaging measures supplementary results

- S3.1 Seasonality associations with brain imaging measures
  - S3.1.1 Global T1 measures
  - S3.1.2 Lobar T1 measures
  - S3.1.3 Individual T1 measures
  - S3.1.4 Subcortical Measures
  - S3.1.5 DTI Global Measures
  - S3.1.6 DTI Grouped Tract Measures
  - S3.1.7 DTI Individual Tract Measures
- S3.2 Seasonality associations with brain imaging measures covarying for birth weight
  - S3.2.1 Global T1 measures
  - S3.2.2 Lobar T1 measures
  - S3.2.3 Individual T1 measures
  - S3.2.4 Subcortical Measures
  - S3.2.5 DTI Global Measures
  - S3.2.6 DTI Grouped Tract Measures
  - S3.2.7 DTI Individual Tract Measures

**SUPPLEMENTARY METHODS**

**S1. UKB Data procedures and acquisition**

***S1.1 Brain Imaging Measures***

Participants were scanned over three locations (Cheadle, Newcastle and Reading) using a Siemens Skyra 3T scanner with a standard Siemens 32-channel RF receive head coil. For this study, T1 and DTI brain imaging measures were extracted from UKB after undergoing a standard pre-processing pipeline [1]. Full protocol and acquisition parameters are available at <https://biobank.ctsu.ox.ac.uk/crystal/crystal/docs/brain_mri.pdf> and <https://www.fmrib.ox.ac.uk/ukbiobank/>.

T1 measures were further processed by UKB with Freesurfer 6.0 software whereby IDPs are extracted in reference to standard surface area, volume and mean cortical thickness given by standard atlases, in this case the Desikan-Killiany-Tourville atlas was utilised [2]. For subcortical regions Freesurfer ASEG was used [3,4]. All output is then subjected to Qoala-T QC checks, with any output close to the threshold also being manually checked [5].

DTI measures are additionally corrected for head motion and eddy currents and fitted with the DTIFIT tool to create fractional anisotropy (FA) and mean diffusivity (MD) outputs. For this study, three tracts were not included in any FA or MD tract group measures (corpus callosum, corona radiata and internal capsule).

**S2. Data pre-processing**

***S2.1 Covariates***

***S2.1.1 Mental health traits***

Variables and UKB data-fields used as covariates used for mental health trait regression models.

| **Covariate** | **UKB data-field** |
| --- | --- |
| Sex | 31 |
| Age | 21003 |
| Age^2^ | - |
| Assessment centre | 54 |
| Townsend Deprivation Index | 189 |
| Birth Location | 129, 130* |

******* Place of birth co-ordinates were collected by UKB as Ordnance Survey grid references <https://biobank.ndph.ox.ac.uk/showcase/showcase/docs/UKgrid.pdf> referring to easting and northing with a reference point close to the Isles of Sicily. Since they adequately tracked north and east directions within the UK in relation to its geography and to each other, they were not converted to true longitude and latitude for this study. Instead, a kmeans clustering approach was utilised whereby participants were clustered by UKB datafields 129 and 130 to derive a birth location cluster after scaling these measures. A maximum of 12 clusters were inspected and the optimal number of clusters was chosen by visual inspection of an elbow chart (See Figures 1-3 below).

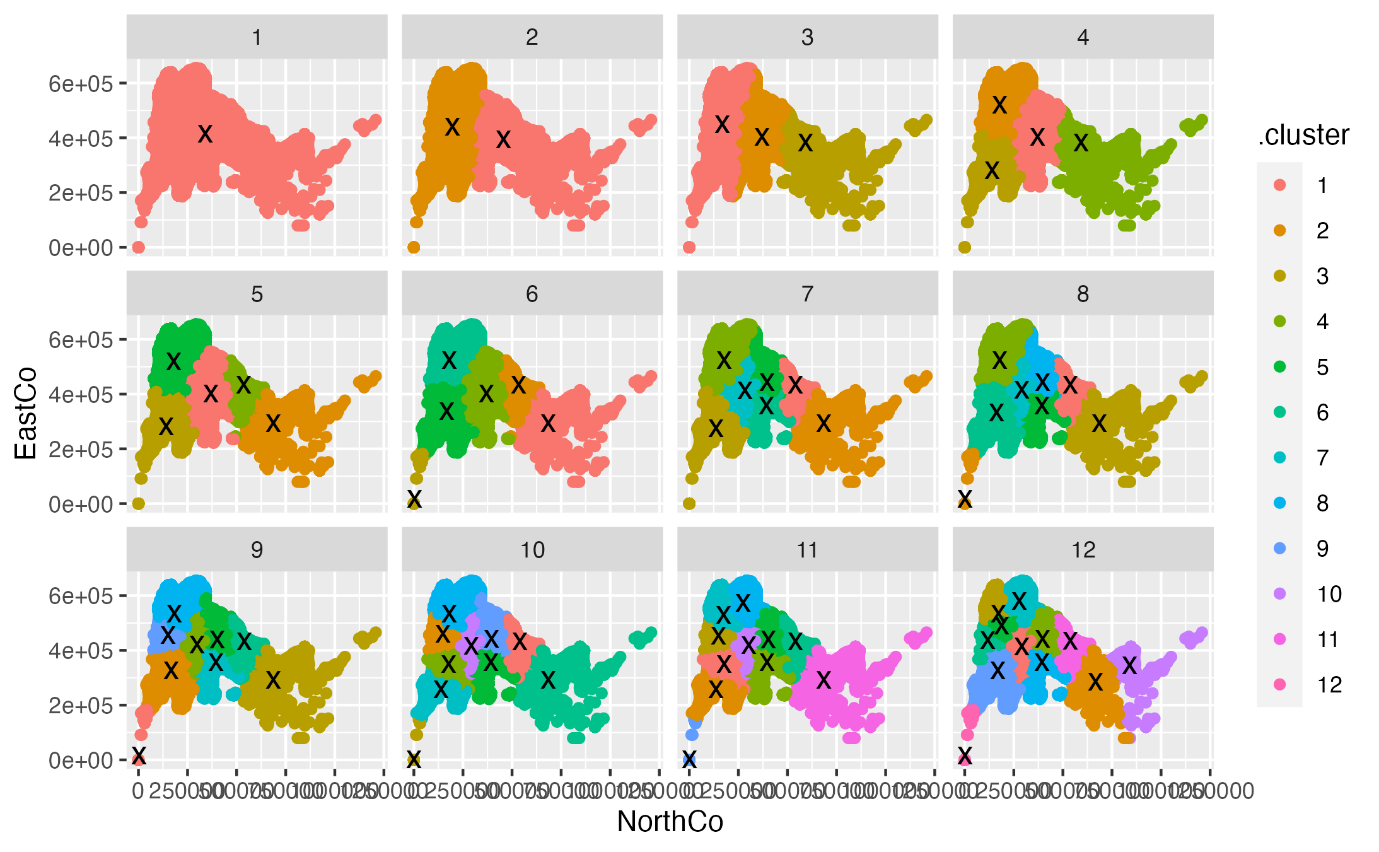

**S1 Fig.** Visualisation of a maximum of 12 clusters for birth location via k-means clustering analysis on UKB datafields 129 and 130 for participants who had completed the MHQ and/or mania and depression questions in the touchscreen questionnaire.
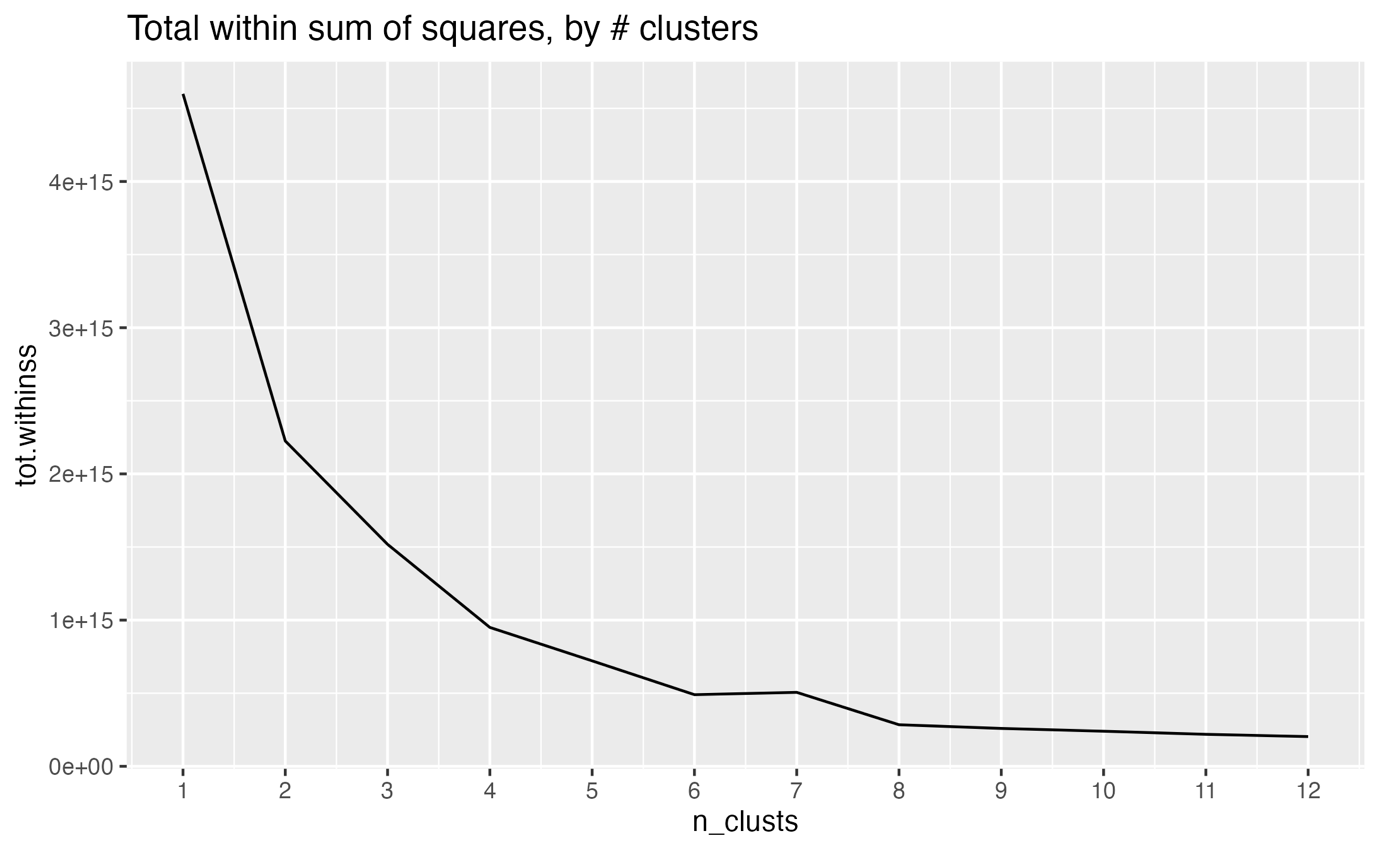

**S2 Fig.** Elbow chart of the total within sum of squares by number of clusters (max N= 12) for kmeans clustering performed on UKB datafields 129 and 130 for participants who had completed the MHQ and/or mania and depression questions in the touchscreen questionnaire.

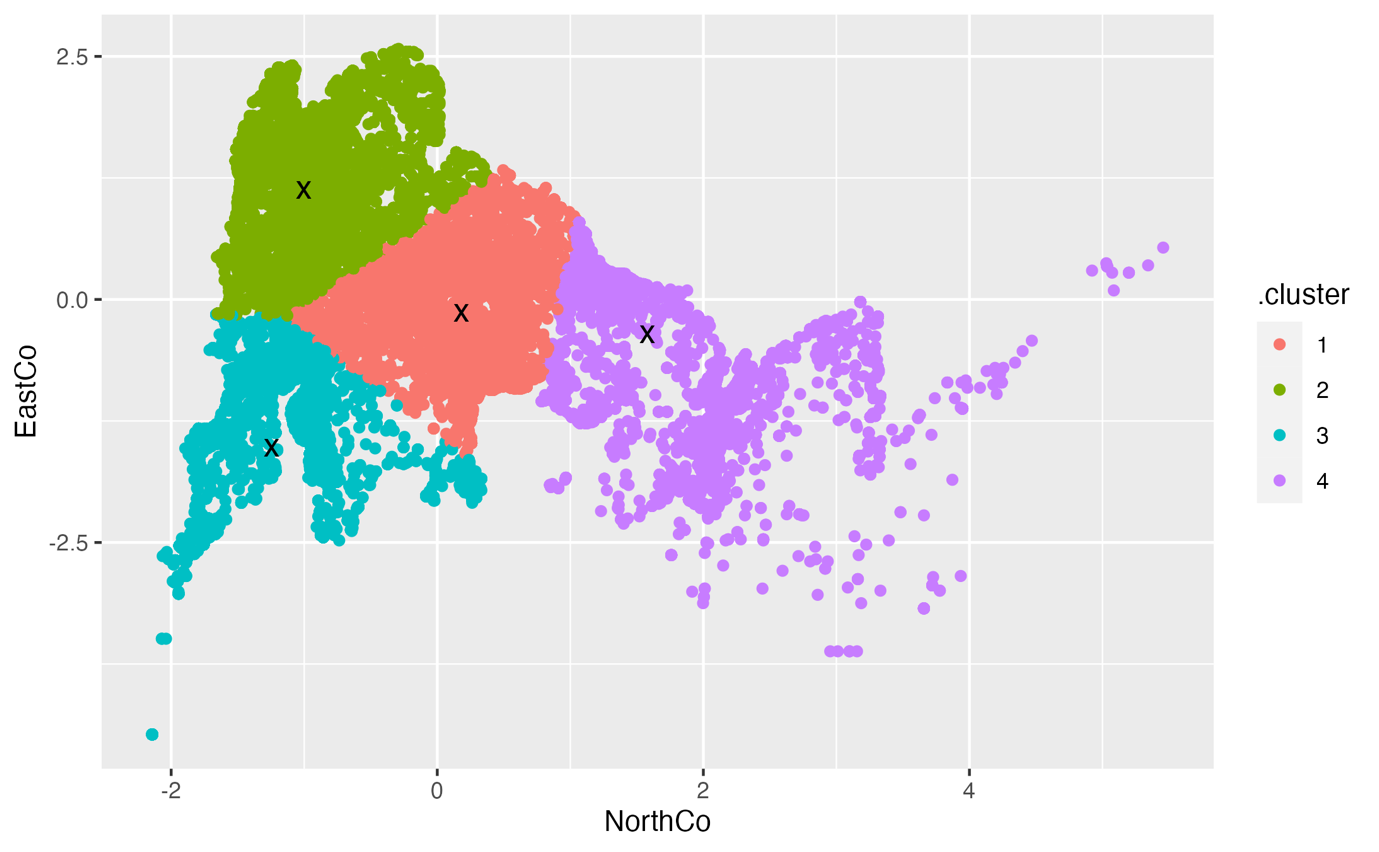

**S3 Fig.** Visualisation of the final four clusters chosen to be the proxy for birth location via k-means clustering analysis on UKB datafields 129 and 130 for participants who had completed the MHQ and/or mania and depression questions in the touchscreen questionnaire.

***S2.1.2 Brain imaging measures***

Variables and UKB data-fields used as covariates used for brain imaging measure regression models.

| **Covariate** | **UKB data-field** |
| --- | --- |
| ***Partially adjusted*** |  |
| Sex | 31 |
| Age | 21003 |
| Age^2^ | - |
| Assessment centre | 54 |
| Standardised intracranial volume | Sum of: 25005, 25007, 25003 |
| Scanner lateral (X) brain position | 25756 |
| Scanner transverse (Y) brain position | 25757 |
| Scanner longitudinal (Z) brain position | 25758 |
| Scanner table position | 25759 |
| Townsend Deprivation Index | 189 |
| Birth Location | 129, 130* |
| ***Maximally adjusted - all of the above plus:*** |  |
| Birth Weight | 20022 |

******* Place of birth co-ordinates were collected by UKB as Ordnance Survey grid references <https://biobank.ndph.ox.ac.uk/showcase/showcase/docs/UKgrid.pdf> referring to easting and northing with a reference point close to the Isles of Sicily. Since they adequately tracked north and east directions within the UK in relation to its geography and to each other, they were not converted to true longitude and latitude for this study. Instead, a kmeans clustering approach was utilised whereby participants were clustered by UKB datafields 129 and 130 to derive a birth location cluster after scaling these measures. A maximum of 12 clusters were inspected and the optimal number of clusters was chosen by visual inspection of an elbow chart (See Figures 4-6 below).

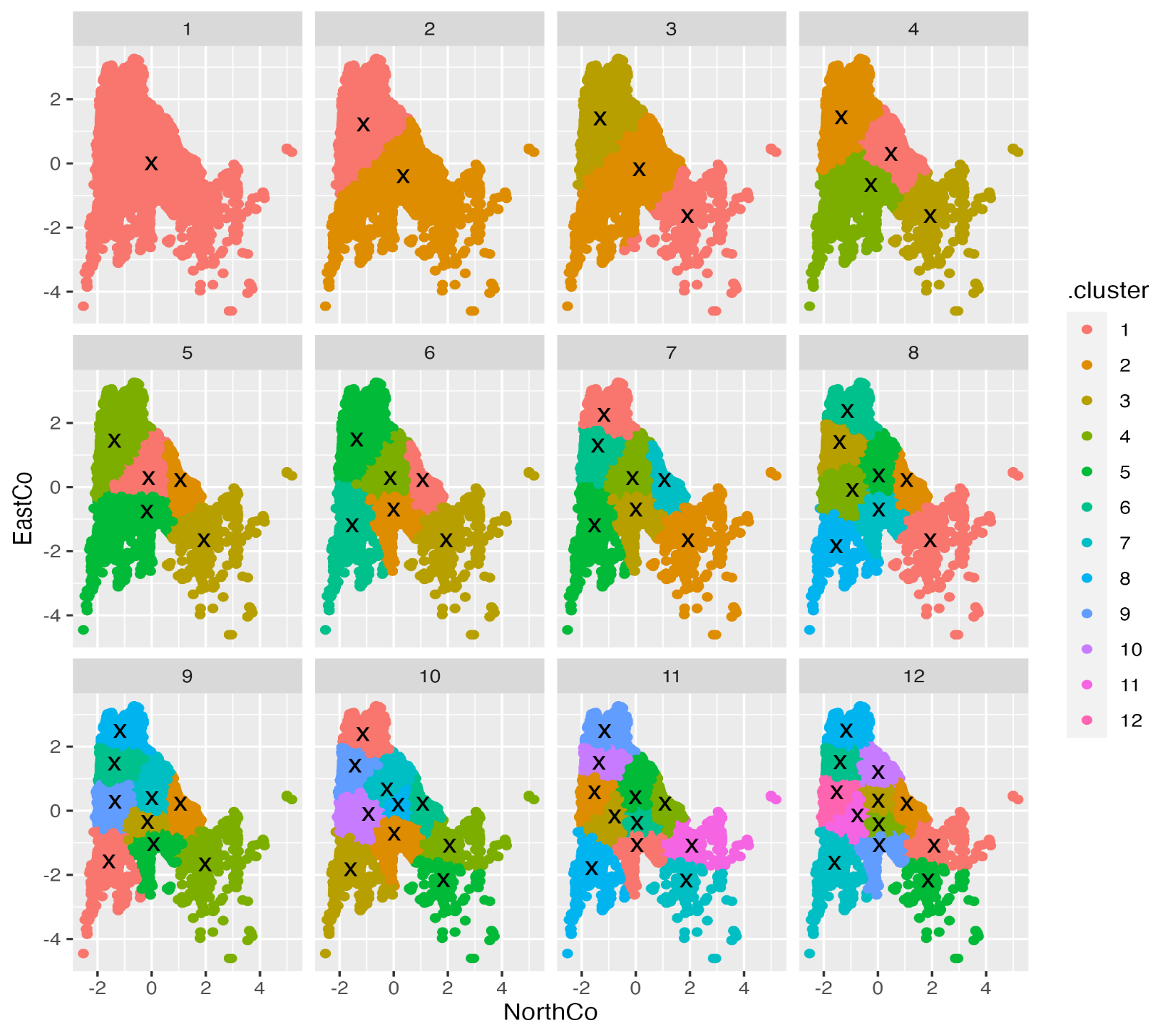
**S4 Fig.** Visualisation of a maximum of 12 clusters for birth location via k-means clustering analysis on UKB datafields 129 and 130 for participants who had attended UKB’s imaging assessment.

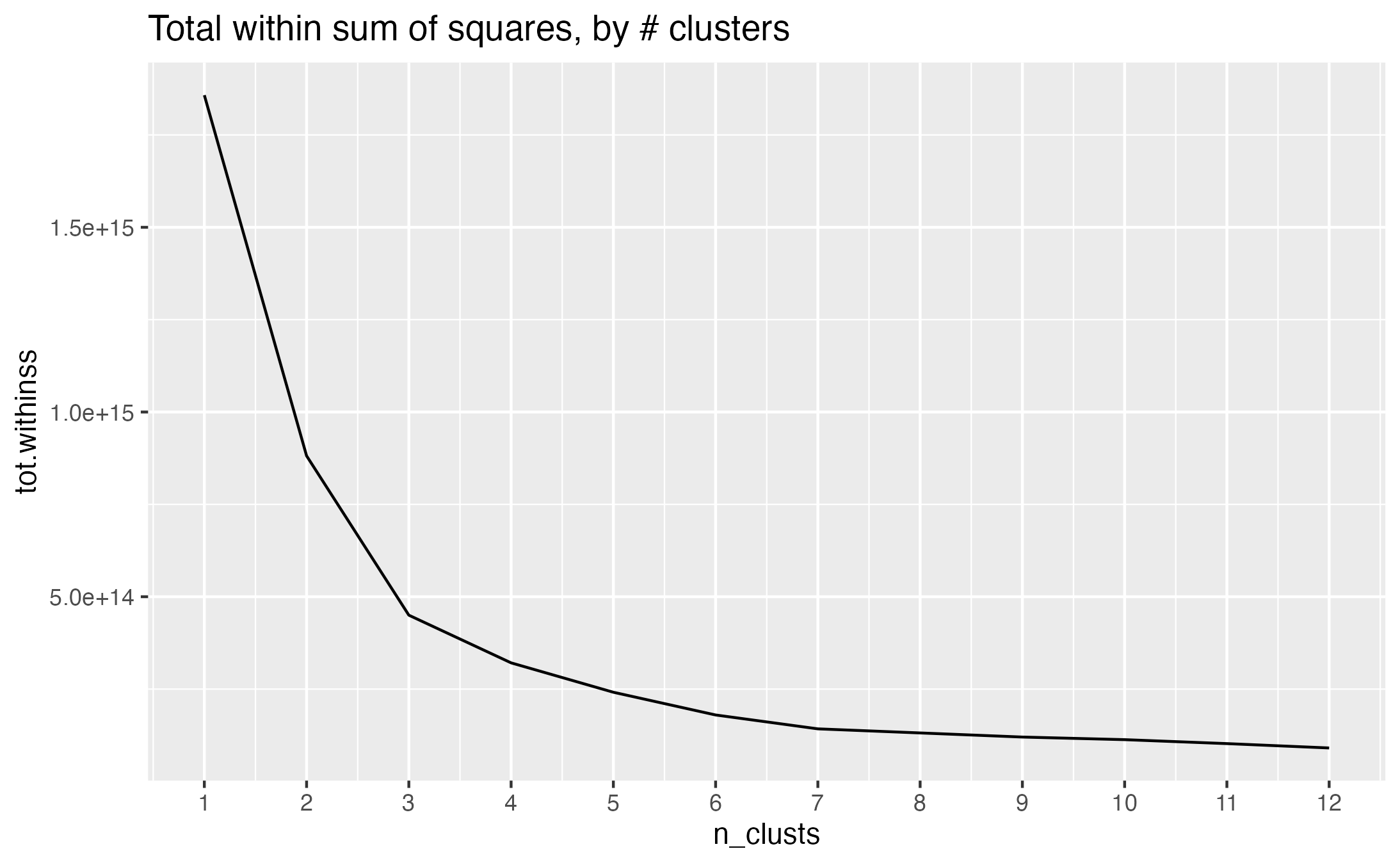
 **S5 Fig.** Elbow chart of the total within sum of squares by number of clusters (max N= 12) for kmeans clustering performed on UKB datafields 129 and 130 for participants who had attended UKB’s imaging assessment.

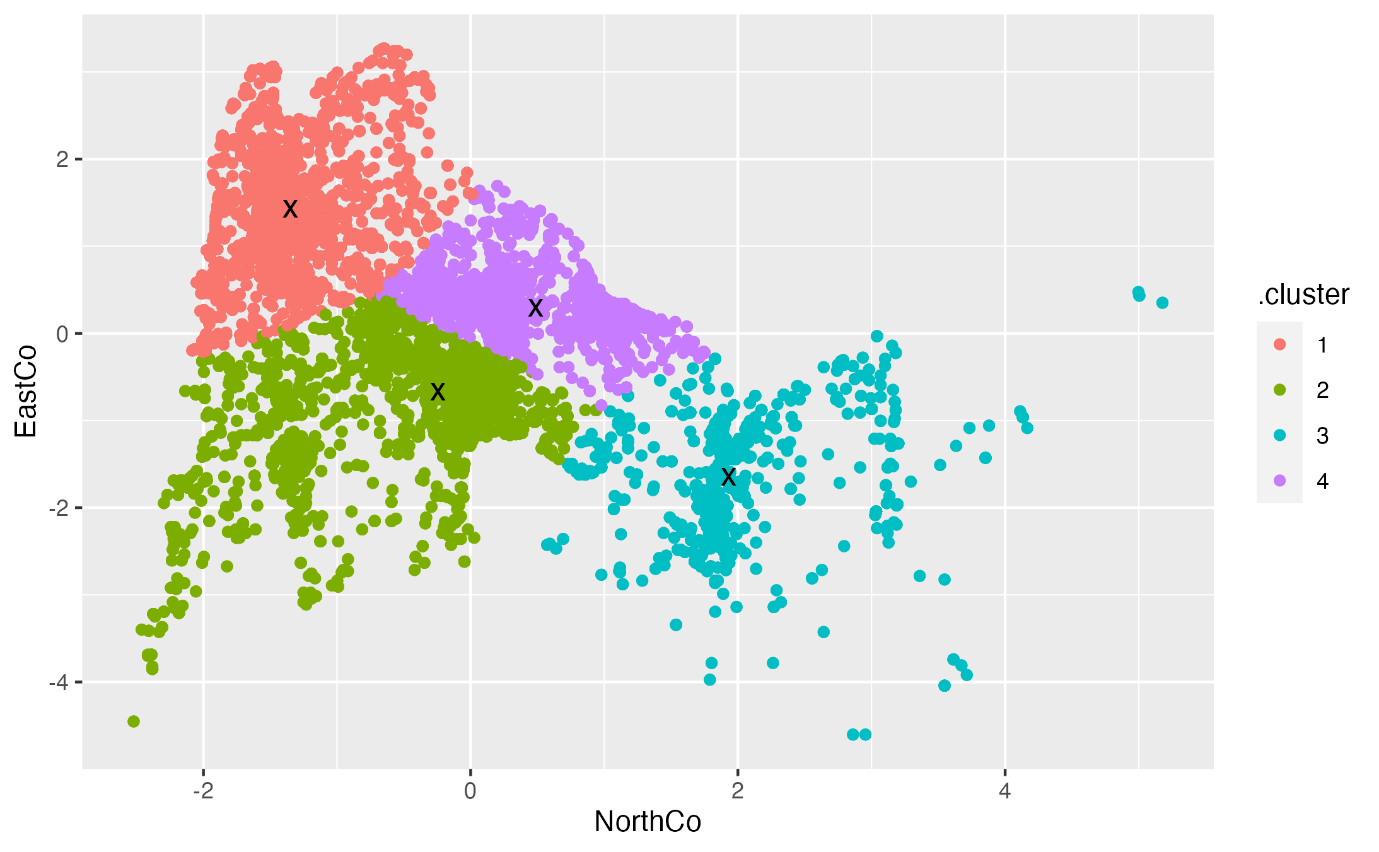

**S6 Fig.** Visualisation of the final four clusters chosen to be the proxy for birth location via k-means clustering analysis on UKB datafields 129 and 130 for participants who had attended UKB’s imaging assessment.

***S2.2 Classification***

***S2.2.1 Mental health traits***

***S2.2.1A Variables utilised to derive probable mental health phenotypes***

UBK variables used to derive probable mania, probable hypomania and no mania (control) mental health trait phenotypes from the Thoughts and Feelings Questionnaire and Touchscreen Mental Health Questionnaire.

|  | **Thoughts and Feelings Questionnaire (MHQ)** | **UKB Data-Fields** | **Touchscreen Mental Health Questionnaire** | **UKB Data-Fields** |
| --- | --- | --- | --- | --- |
| ***Probable Mania*** | Yes to hyper/manic for two days  OR | 20501 | Yes to hyper/manic for two days  OR | 4642 |
|  | Yes to irritable/argumentative for two days | 20502 | Yes to irritable/argumentative for two days | 4653 |
|  | Three or more symptoms | 20548 | Three or more symptoms | 6156 |
|  | Episode length of one week or more | 20492 | Episode length of one week or more | 5663 |
|  | Symptoms affected daily activities | 20493 | Symptoms affected daily activities | 5674 |
| ***Probable Hypomania*** | Yes to hyper/manic for two days  OR | 20501 | Yes to hyper/manic for two days  OR | 4642 |
|  | Yes to irritable/argumentative for two days | 20502 | Yes to irritable/argumentative for two days | 4653 |
|  | Three or more symptoms | 20548 | Three or more symptoms | 6156 |
|  | Episode length of 24 hours or more | 20492 | Episode length of two days or more | 5663 |
| ***No Mania*** | No to hyper/manic for two days  AND | 20501 | No to hyper/manic for two days  AND | 4642 |
|  | No to irritable/argumentative for two days | 20502 | No to irritable/argumentative for two days | 4653 |

***S2.2.1B Variables utilised to derive probable depression mental health phenotypes***

UKB variables utilised to derive probable singular episode of major depression, probable recurrent depression and no probable depression (control) mental health trait phenotypes from the Thoughts and Feelings Questionnaire and Touchscreen Mental Health Questionnaire.

| **Phenotype** | **Thoughts and Feelings Questionnaire (MHQ)** | **UKB Data-Fields** | **Touchscreen Mental Health Questionnaire** | **UKB Data-Fields** |
| --- | --- | --- | --- | --- |
| ***Probable Single Episode of Major Depression*** | Yes to depressed feelings for two+ weeks  OR | 20446 | Yes to depressed feelings for one week  OR | 4598 |
|  | Yes to anhedonia for two+ weeks | 20441 | Yes to anhedonia for one week | 4631 |
|  | Five or more symptoms | 20446, 20441, 20536, 20532, 20450, 20435, 20437 | Maximum period of depression/anhedonia lasted two+ weeks | 4609 |
|  | One episode over lifetime | 20442 | One episode over lifetime | 4620 |
|  | Professional informed about depression | 20448 | GP or psychiatrist seen for nerves/anxiety/tension/depression | 2090, 2100 |
| ***Probable Recurrent Depression*** | Yes to depressed feelings for two+ weeks  OR | 20446 | Yes to depressed feelings for one week  OR | 4598 |
|  | Yes to anhedonia for two+ weeks | 20441 | Yes to anhedonia for one week | 4631 |
|  | Five or more symptoms | 20446, 20441, 20536, 20532, 20450, 20435, 20437 | Maximum period of depression/anhedonia lasted two+ weeks | 4609 |
|  | More than one episode over lifetime | 20442 | More than one episode over lifetime | 4620 |
|  | Professional informed about depression | 20448 | GP or psychiatrist seen for nerves/anxiety/tension/depression | 2090, 2100 |
| ***No Probable Depression*** | No to depressed feelings for two+ weeks AND | 20446 | No to depressed feelings for one week AND | 4598 |
|  | No to anhedonia for two+ weeks | 20441 | No to anhedonia for one week | 4631 |

***S2.2.1C Criteria for mental health trait phenotype groupings***

Criteria used to define final mental health trait phenotype groupings based on participant response type to questions from UKB Thoughts and Feelings Questionnaire and UKB Touchscreen Mental Health Questionnaire.

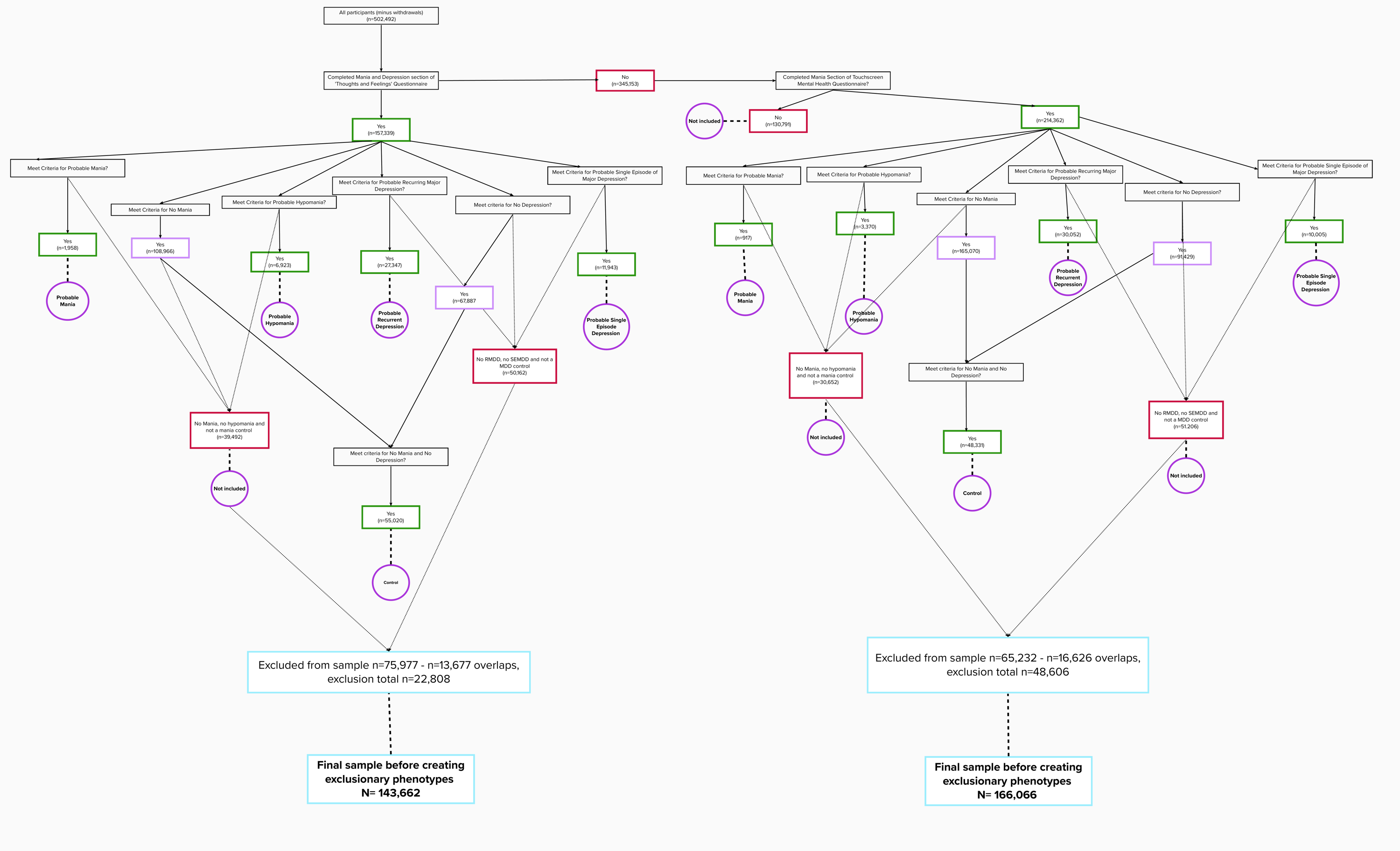

***S2.2.1D Overlaps in mental health phenotype groupings***

Overlaps permitted within the final mental health phenotype groupings. All participants in the control group had answered ‘no’ to leading probable mania and/or probable depression.

|  | **Probable Unipolar Mania** |  |  |  | **Probable Bipolar Depression** |  | **Probable Recurring MDD** | **Probable Single Episode MDD** | **Control Group** |
| --- | --- | --- | --- | --- | --- | --- | --- | --- | --- |
| **Probable Mania** | Y | N | Y | Y | N | N | N | N | N |
| **Probable Hypomania** | N | Y | N | N | Y | Y | N | N | N |
| **Probable Recurring MDD** | N | N | Y | N | Y | N | Y | N | N |
| **Probable Single Episode MDD** | N | N | N | Y | N | Y | N | Y | N |

***S2.2.2 Brain imaging measures***

***S2.2.2A UKB T1 and DTI brain imaging variables used to derive brain imaging measures***

UKB variables used to derive brain imaging measures per participant. Individual T1 measures per hemisphere were extracted for each participant from the Freesurfer DKT category (196) for bilateral measures and unilaterally for unilateral measures. Lobar and global T1 measures were composed per participant as below. Subcortical structures were extracted from the UKB ‘FIRST’ category (1102) by hemisphere. FA and MD DTI measures were extracted per hemisphere for bilateral structures from the ‘dMRI weighted means’ UKB category (135) and unilaterally for unilateral measures.

| **Variable** | **Measures** |
| --- | --- |
| ***T1 Global Measures*** |  |
| Global Cortical Volume | Sum of 5 lobar measures |
| Global Cortical Thickness | Weighted average of thickness of the 5 lobes multiplied by their surface area and divided by their thickness. |
| Global Surface Area | Sum of 5 lobar measures |
| ***T1 Lobes*** |  |
| Frontal | Sum of: Superior frontal gyrus, Rostral middle frontal, caudal middle frontal, Pars orbitalis, pars triangularis, pars opercularis, Lateral orbitofrontal, medial orbitofrontal, Precentral gyrus, Paracentral cortex |
| Temporal | Sum of: Insula, Superior temporal, transverse temporal, Middle temporal gyrus, Inferior temporal gyrus, Fusiform, parahippocampal, entorhinal |
| Parietal | Sum of: Postcentral gyrus, paracentral cortex, Superior parietal cortex, Inferior parietal cortex, Supramarginal gyrus, Precuneus |
| Occipital | Sum of: Lateral occipital cortex, Cuneus, Pericalcarine cortex, Lingual gyrus |
| Cingulate | Sum of: Rostral anterior cingulate cortex, Caudal anterior cingulate cortex, Posterior cingulate cortex, Cingulate isthmus |
| ***T1 Individual Structures*** |  |
| 31 Cortical Regions | All of the above in the lobe categories |
| 7 Subcortical | Nucelus accumbens, amygdala, caudate nucleus, hippocampus, pallidum, putamen and thalamus |
| ***DTI Measures*** |  |
| gFA/MD | PCA of all tracts |
| gAssociation Fibres FA/MD | PCA of 6 bilateral tracts; Cingulum-Cingulate Gyrus, Parahippocampal part of cingulum, inferior fronto-occipital-fasciculus, superior longitudinal fasciculus, uncinate fasciculus, inferior longitudinal fasciculus). |
| gProjection Fibres FA/MD | PCA of 6 bilateral tracts (3 bilateral, 3 unilateral); Forceps major, forceps minor, corticospinal tract, acoustic radiation, medial lemniscus, middle cerebellar peduncle. |
| gThalamic Radiations FA/MD | PCA of 3 bilateral tracts; Superior thalamic radiation, posterior thalamic radiation, anterior thalamic radiation |
| Individual Tracts | All of the above individually. |

***S2.2.2B Variance explained by the first principal component for DTI PCA***

For each global and grouped DTI FA and MD tract measure, the variance explained by the first principal component.

| **PCA** | **FA** | **MD** |
| --- | --- | --- |
| gTotal | 37.1% | 37.6% |
| Association Fibres | 44.6% | 50% |
| Projection Fibres | 35.3% | 29.5% |
| Thalamic Radiations | 61.0% | 71.5% |

***S2.3 Quality Control
S2.3.1 Mental health traits***

***S2.3.1A Conditions excluded for mental health traits***
Conditions excluded per participant for final mental health trait phenotypes and data-field the4y correspond to. Response variable corresponds to UKB data coding.

| **Exclusion Type** | **Response variable** |
| --- | --- |
| ***Neuropsychiatric conditions (UKB Data-Field 20002)*** |  |
| Stroke | 1081 |
| Transient ischaemic attack | 1082 |
| Subdural haemorrhage/ Haematoma | 1083 |
| Subarachnoid haemorrhage | 1086 |
| Neurological injury/trauma | 1240 |
| Infection of nervous system | 1244 |
| Brain abscess/Intracranial abscess | 1245 |
| Meningitis | 1247 |
| Chronic/degenerative neurological problem | 1258 |
| Motor neurone disease | 1259 |
| Multiple sclerosis | 1261 |
| Dementia/Alzheimers/Cognitive impairment | 1263 |
| Migraine | 1265 |
| Head injury | 1266 |
| Other demyelinating disease | 1397 |
| Cerebral aneurysm | 1425 |
| Cerebral palsy | 1433 |
| Other neurological problem | 1434 |
| Brain haemorrhage | 1491 |
| Spina bifida | 1524 |
| Ischaemic stroke | 1583 |
| Fracture skull / head | 1626 |
| Meningioma / Benign meningeal tumour | 1659 |
| Benign neuroma | 1683 |
| ***Sleep conditions (UKB Data-Field 20002)*** |  |
| Insomnia | 1616 |
| Sleep apnoea | 1123 |
| ***Cancer (UKB Data-Field 20001)*** |  |
| Meningeal cancer / malignant meningioma | 1032 |
| Brain cancer / primary malignant brain tumour | 1031 |
| ***Shift Work (UKB Data-Field 826)*** |  |
| Job involving shift work | 2, 3 |

***S2.3.1B Sample size per mental health traits post-exclusions***

Sample size for each (*n*=4) mental health trait phenotype after grouping and exclusions.

| **Mental Health Trait** | **Total N** |
| --- | --- |
| Probable Recurrent MDD | 31,652 |
| Probable Single Episode MDD | 13,529 |
| Unipolar Mania | 1,202 |
| Bipolar Depression | 5,140 |
| Control | 84,018 |

***S2.4 Multiple testing correction***

***S2.4.1 Multiple testing corrections applied per neuroimaging measure modality***
Number of neuroimaging measures False Discovery Rate (FDR) was applied for.

| **Measure name** | **Number of measures FDR was applied for** | **Measure name** | **Number of measures FDR was applied for** |
| --- | --- | --- | --- |
| **Bilateral** |  | **Unilateral** |  |
| Individual Cortical Area | 31 | Global Cortical Area |  |
| Individual Mean Cortical Thickness | 31 | Global Mean Cortical Thickness |  |
| Individual Cortical Volume | 31 | Global Cortical Volume |  |
|  | | Lobar Cortical Area | 5 |
|  |  | Lobar Cortical Thickness | 5 |
|  |  | Lobar Cortical Volume | 5 |
| Individual Subcortical Volume | 7 |  | |
| FA Individual | 15 | Global FA |  |
| MD Individual | 15 | Global MD |  |
|  | | FA tract bundles | 3 |
|  |  | MD tract bundles | 3 |

**SUPPLEMENTARY RESULTS**

***S3. Brain Imaging Measures Supplementary Results***

***S3.1.1 Global T1 measures***

Global T1 brain measure associations with Seasonality.

p-uncorr., p-uncorrected value; p-corr., FDR p-corrected value; S.E., standard error.

| **Brain Imaging Measure** | **Effect Size (β)** | **S.E.** | **t statistic** | **p-uncorr.** | **p-corr** |
| --- | --- | --- | --- | --- | --- |
| GlobalSurfaceArea | -0.007 | 0.004 | -1.839 | 0.066 | - |
| GlobalCorticalVolume | -0.001 | 0.004 | -0.347 | 0.729 | - |
| GlobalCorticalThickness | -0.006 | 0.004 | -1.415 | 0.157 | - |

***S3.1.2 Lobar T1 measures***

Lobar T1 brain measure associations with Seasonality.

p-uncorr., p-uncorrected value; p-corr., FDR p-corrected value; S.E., standard error.

| **Brain Imaging Measure** | **Effect Size (β)** | **S.E.** | **t statistic** | **p-uncorr.** | **p-corr** |
| --- | --- | --- | --- | --- | --- |
| ***Area*** |  |  |  |  |  |
| FrontalArea | -0.008 | 0.003 | -2.356 | 0.018 | 0.070 |
| CingulateArea | -0.007 | 0.003 | -2.200 | 0.028 | 0.070 |
| OccipitalArea | -0.006 | 0.003 | -1.732 | 0.083 | 0.139 |
| TemporalArea | -0.002 | 0.003 | -0.689 | 0.491 | 0.491 |
| ParietalArea | 0.003 | 0.003 | 0.764 | 0.445 | 0.491 |
| ***Volume*** |  |  |  |  |  |
| FrontalVolume | -0.007 | 0.003 | -2.276 | 0.023 | 0.114 |
| CingulateVolume | -0.006 | 0.003 | -1.831 | 0.067 | 0.168 |
| TemporalVolume | 0.002 | 0.003 | 0.688 | 0.491 | 0.614 |
| OccipitalVolume | -0.003 | 0.003 | -0.720 | 0.472 | 0.614 |
| ParietalVolume | -0.001 | 0.003 | -0.219 | 0.827 | 0.827 |
| ***Thickness*** |  |  |  |  |  |
| **TemporalThickness** | **0.011** | **0.004** | **2.922** | **0.003** | **0.017** |
| **OccipitalThickness** | **0.010** | **0.004** | **2.569** | **0.010** | **0.025** |
| CingulateThickness | 0.007 | 0.004 | 1.736 | 0.083 | 0.138 |
| ParietalThickness | -0.004 | 0.004 | -1.187 | 0.235 | 0.294 |
| FrontalThickness | 0.000 | 0.004 | -0.023 | 0.982 | 0.982 |

***S3.1.3 Individual T1 measures***

Individual T1 brain measure associations with Seasonality.

p-uncorr., p-uncorrected value; p-corr., FDR p-corrected value; S.E., standard error; DF., degrees of freedom

| **Brain Imaging Measure** | **Effect Size (β)** | **S.E.** | **DF** | **t statistic** | **p-uncorr.** | **p-corr** |
| --- | --- | --- | --- | --- | --- | --- |
| ***Surface area*** |  |  |  |  |  |  |
| Areaofcaudalanteriorcingulate | -0.012 | 0.004 | 21166.000 | -3.148 | 0.002 | 0.051 |
| Areaofparacentral | -0.014 | 0.006 | 21166.000 | -2.544 | 0.011 | 0.170 |
| Areaofcuneus | -0.013 | 0.006 | 21166.000 | -2.196 | 0.028 | 0.203 |
| Areaofpericalcarine | -0.013 | 0.006 | 21166.000 | -2.136 | 0.033 | 0.203 |
| Areaofsuperiorfrontal | -0.011 | 0.005 | 21166.000 | -2.244 | 0.025 | 0.203 |
| Areaofprecentral | -0.011 | 0.006 | 21166.000 | -2.058 | 0.040 | 0.205 |
| Areaofmedialorbitofrontal | -0.010 | 0.005 | 21166.000 | -1.886 | 0.059 | 0.227 |
| Areaofparahippocampal | -0.010 | 0.006 | 21166.000 | -1.885 | 0.060 | 0.227 |
| Areaoftransversetemporal | -0.007 | 0.004 | 21166.000 | -1.838 | 0.066 | 0.227 |
| Areaofposteriorcingulate | -0.009 | 0.005 | 21166.000 | -1.696 | 0.090 | 0.279 |
| Areaofparsorbitalis | -0.009 | 0.005 | 21166.000 | -1.597 | 0.110 | 0.310 |
| Areaofcaudalmiddlefrontal | -0.009 | 0.006 | 21166.000 | -1.541 | 0.123 | 0.319 |
| Areaofinsula | -0.008 | 0.005 | 21166.000 | -1.454 | 0.146 | 0.348 |
| Areaoflateraloccipital | -0.007 | 0.006 | 21166.000 | -1.342 | 0.180 | 0.371 |
| Areaofsuperiortemporal | -0.007 | 0.005 | 21166.000 | -1.350 | 0.177 | 0.371 |
| Areaoflingual | -0.007 | 0.006 | 21166.000 | -1.245 | 0.213 | 0.413 |
| Areaoffusiform | -0.006 | 0.005 | 21166.000 | -1.105 | 0.269 | 0.464 |
| Areaofparsopercularis | 0.006 | 0.005 | 21166.000 | 1.119 | 0.263 | 0.464 |
| Areaofentorhinal | 0.004 | 0.006 | 21166.000 | 0.769 | 0.442 | 0.689 |
| Areaofrostralanteriorcingulate | -0.003 | 0.004 | 21166.000 | -0.728 | 0.467 | 0.689 |
| Areaofrostralmiddlefrontal | -0.004 | 0.006 | 21166.000 | -0.760 | 0.447 | 0.689 |
| Areaofmiddletemporal | 0.004 | 0.006 | 21166.000 | 0.658 | 0.511 | 0.720 |
| Areaofinferiortemporal | 0.003 | 0.005 | 21166.000 | 0.481 | 0.631 | 0.751 |
| Areaoflateralorbitofrontal | -0.002 | 0.006 | 21166.000 | -0.448 | 0.654 | 0.751 |
| Areaofparstriangularis | -0.003 | 0.005 | 21166.000 | -0.573 | 0.567 | 0.751 |
| Areaofpostcentral | -0.003 | 0.005 | 21166.000 | -0.494 | 0.621 | 0.751 |
| Areaofsupramarginal | 0.003 | 0.005 | 21166.000 | 0.472 | 0.637 | 0.751 |
| Areaofsuperiorparietal | -0.002 | 0.006 | 21166.000 | -0.317 | 0.751 | 0.832 |
| Areaofprecuneus | 0.001 | 0.005 | 21166.000 | 0.205 | 0.838 | 0.896 |
| Areaofinferiorparietal | 0.001 | 0.005 | 21166.000 | 0.117 | 0.907 | 0.907 |
| Areaofisthmuscingulate | 0.001 | 0.005 | 21166.000 | 0.123 | 0.902 | 0.907 |
| ***Thickness*** |  |  |  |  |  |  |
| **Meanthicknessofmiddletemporal** | **0.018** | **0.005** | **21166.000** | **3.326** | **0.001** | **0.027** |
| **Meanthicknessoffusiform** | **0.018** | **0.006** | **21166.000** | **3.094** | **0.002** | **0.031** |
| **Meanthicknessofsuperiortemporal** | **0.016** | **0.006** | **21166.000** | **2.944** | **0.003** | **0.033** |
| **Meanthicknessoflingual** | **0.016** | **0.006** | **21166.000** | **2.729** | **0.006** | **0.049** |
| Meanthicknessofcaudalanteriorcingulate | 0.012 | 0.005 | 21166.000 | 2.333 | 0.020 | 0.087 |
| Meanthicknessofcuneus | 0.014 | 0.006 | 21166.000 | 2.431 | 0.015 | 0.087 |
| Meanthicknessofparahippocampal | 0.014 | 0.006 | 21166.000 | 2.385 | 0.017 | 0.087 |
| Meanthicknessofinferiortemporal | 0.013 | 0.006 | 21166.000 | 2.216 | 0.027 | 0.092 |
| Meanthicknessofpericalcarine | 0.013 | 0.006 | 21166.000 | 2.248 | 0.025 | 0.092 |
| Meanthicknessofrostralanteriorcingulate | 0.011 | 0.005 | 21166.000 | 2.072 | 0.038 | 0.119 |
| Meanthicknessoflateraloccipital | 0.011 | 0.006 | 21166.000 | 1.878 | 0.060 | 0.170 |
| Meanthicknessofmedialorbitofrontal | 0.009 | 0.006 | 21166.000 | 1.574 | 0.116 | 0.298 |
| Meanthicknessofentorhinal | 0.008 | 0.006 | 21166.000 | 1.383 | 0.167 | 0.309 |
| Meanthicknessofinsula | 0.008 | 0.006 | 21166.000 | 1.424 | 0.154 | 0.309 |
| Meanthicknessofisthmuscingulate | 0.008 | 0.006 | 21166.000 | 1.375 | 0.169 | 0.309 |
| Meanthicknessofparacentral | 0.008 | 0.006 | 21166.000 | 1.410 | 0.159 | 0.309 |
| Meanthicknessoftransversetemporal | 0.008 | 0.006 | 21166.000 | 1.406 | 0.160 | 0.309 |
| Meanthicknessofparsorbitalis | 0.005 | 0.006 | 21166.000 | 0.951 | 0.342 | 0.588 |
| Meanthicknessofinferiorparietal | 0.004 | 0.006 | 21166.000 | 0.740 | 0.459 | 0.735 |
| Meanthicknessofparstriangularis | 0.004 | 0.006 | 21166.000 | 0.715 | 0.474 | 0.735 |
| Meanthicknessofcaudalmiddlefrontal | 0.003 | 0.006 | 21166.000 | 0.547 | 0.584 | 0.791 |
| Meanthicknessoflateralorbitofrontal | 0.003 | 0.006 | 21166.000 | 0.574 | 0.566 | 0.791 |
| Meanthicknessofrostralmiddlefrontal | 0.003 | 0.006 | 21166.000 | 0.515 | 0.607 | 0.791 |
| Meanthicknessofsuperiorfrontal | 0.003 | 0.006 | 21166.000 | 0.507 | 0.612 | 0.791 |
| Meanthicknessofprecentral | 0.002 | 0.006 | 21166.000 | 0.396 | 0.692 | 0.825 |
| Meanthicknessofsupramarginal | 0.002 | 0.006 | 21166.000 | 0.398 | 0.691 | 0.825 |
| Meanthicknessofposteriorcingulate | -0.002 | 0.005 | 21166.000 | -0.314 | 0.754 | 0.856 |
| Meanthicknessofprecuneus | -0.002 | 0.006 | 21166.000 | -0.288 | 0.773 | 0.856 |
| Meanthicknessofsuperiorparietal | -0.001 | 0.006 | 21166.000 | -0.250 | 0.803 | 0.858 |
| Meanthicknessofpostcentral | -0.001 | 0.006 | 21166.000 | -0.098 | 0.922 | 0.953 |
| Meanthicknessofparsopercularis | 0.000 | 0.006 | 21166.000 | 0.003 | 0.997 | 0.997 |
| ***Volume*** |  |  |  |  |  |  |
| Volumeofcaudalanteriorcingulate | -0.006 | 0.004 | 21166.000 | -1.494 | 0.135 | 0.865 |
| Volumeofcaudalmiddlefrontal | -0.007 | 0.006 | 21166.000 | -1.244 | 0.213 | 0.865 |
| Volumeofentorhinal | 0.007 | 0.006 | 21166.000 | 1.322 | 0.186 | 0.865 |
| Volumeofinferiortemporal | 0.007 | 0.005 | 21166.000 | 1.218 | 0.223 | 0.865 |
| Volumeofmiddletemporal | 0.009 | 0.005 | 21166.000 | 1.669 | 0.095 | 0.865 |
| Volumeofparacentral | -0.007 | 0.006 | 21166.000 | -1.288 | 0.198 | 0.865 |
| Volumeofposteriorcingulate | -0.010 | 0.005 | 21166.000 | -1.929 | 0.054 | 0.865 |
| Volumeofsuperiorfrontal | -0.009 | 0.005 | 21166.000 | -1.811 | 0.070 | 0.865 |
| Volumeofisthmuscingulate | 0.006 | 0.005 | 21166.000 | 1.054 | 0.292 | 0.896 |
| Volumeofmedialorbitofrontal | -0.005 | 0.005 | 21166.000 | -0.937 | 0.349 | 0.896 |
| Volumeofparsopercularis | 0.005 | 0.005 | 21166.000 | 0.967 | 0.334 | 0.896 |
| Volumeofpericalcarine | -0.005 | 0.006 | 21166.000 | -0.885 | 0.376 | 0.896 |
| Volumeofprecentral | -0.006 | 0.006 | 21166.000 | -1.033 | 0.301 | 0.896 |
| Volumeofcuneus | -0.002 | 0.006 | 21166.000 | -0.320 | 0.749 | 0.969 |
| Volumeoffusiform | 0.001 | 0.005 | 21166.000 | 0.205 | 0.838 | 0.969 |
| Volumeofinferiorparietal | 0.002 | 0.005 | 21166.000 | 0.417 | 0.677 | 0.969 |
| Volumeofinsula | -0.002 | 0.006 | 21166.000 | -0.298 | 0.766 | 0.969 |
| Volumeoflateraloccipital | -0.002 | 0.005 | 21166.000 | -0.380 | 0.704 | 0.969 |
| Volumeoflingual | 0.002 | 0.006 | 21166.000 | 0.314 | 0.753 | 0.969 |
| Volumeofparahippocampal | 0.000 | 0.006 | 21166.000 | 0.078 | 0.938 | 0.969 |
| Volumeofparsorbitalis | -0.002 | 0.005 | 21166.000 | -0.377 | 0.706 | 0.969 |
| Volumeofparstriangularis | -0.001 | 0.005 | 21166.000 | -0.120 | 0.904 | 0.969 |
| Volumeofpostcentral | -0.001 | 0.005 | 21166.000 | -0.228 | 0.820 | 0.969 |
| Volumeofprecuneus | 0.000 | 0.006 | 21166.000 | -0.083 | 0.934 | 0.969 |
| Volumeofrostralanteriorcingulate | 0.000 | 0.004 | 21166.000 | -0.102 | 0.919 | 0.969 |
| Volumeofrostralmiddlefrontal | -0.002 | 0.006 | 21166.000 | -0.382 | 0.702 | 0.969 |
| Volumeofsuperiorparietal | -0.003 | 0.006 | 21166.000 | -0.455 | 0.649 | 0.969 |
| Volumeofsuperiortemporal | 0.002 | 0.005 | 21166.000 | 0.441 | 0.659 | 0.969 |
| Volumeofsupramarginal | 0.004 | 0.005 | 21166.000 | 0.698 | 0.485 | 0.969 |
| Volumeoftransversetemporal | -0.001 | 0.005 | 21166.000 | -0.269 | 0.788 | 0.969 |
| Volumeoflateralorbitofrontal | 0.000 | 0.006 | 21166.000 | 0.007 | 0.994 | 0.994 |

***S3.1.4 Subcortical Measures***

Subcortical brain measure associations with Seasonality.

p-uncorr., p-uncorrected value; p-corr., FDR p-corrected value; S.E., standard error; DF., degrees of freedom

| **Brain Imaging Measure** | **Effect Size (β)** | **S.E.** | **DF** | **t statistic** | **p-uncorr.** | **p-corr** |
| --- | --- | --- | --- | --- | --- | --- |
| ***Volume*** |  |  |  |  |  |  |
| VolumeOfamygdala | 0.013 | 0.005 | 21166.000 | 2.424 | 0.015 | 0.108 |
| VolumeOfaccumbens | -0.005 | 0.005 | 21166.000 | -1.063 | 0.288 | 0.672 |
| VolumeOfthalamus | -0.006 | 0.006 | 21166.000 | -1.098 | 0.272 | 0.672 |
| VolumeOfcaudate | 0.003 | 0.006 | 21166.000 | 0.483 | 0.629 | 0.898 |
| VolumeOfhippocampus | 0.001 | 0.006 | 21166.000 | 0.187 | 0.852 | 0.898 |
| VolumeOfpallidum | 0.001 | 0.006 | 21166.000 | 0.140 | 0.889 | 0.898 |
| VolumeOfputamen | 0.001 | 0.006 | 21166.000 | 0.128 | 0.898 | 0.898 |

***S3.1.5 DTI Global Measures***

Global DTI brain measure associations with Seasonality.

p-uncorr., p-uncorrected value; p-corr., FDR p-corrected value; S.E., standard error.

| **Brain Imaging Measure** | **Effect Size (β)** | **S.E.** | **t statistic** | **p-uncorr.** | **p-corr** |
| --- | --- | --- | --- | --- | --- |
| **FATotalTracts** | **-0.013** | **0.004** | **-3.433** | **0.001** | **-** |
| MDTotalTracts | -0.001 | 0.003 | -0.187 | 0.851 | - |

***S3.1.6 DTI Grouped Tract Measures***

Grouped DTI tract brain measure associations with Seasonality.

p-uncorr., p-uncorrected value; p-corr., FDR p-corrected value; S.E., standard error.

| **Brain Imaging Measure** | **Effect Size (β)** | **S.E.** | **t statistic** | **p-uncorr.** | **p-corr** |
| --- | --- | --- | --- | --- | --- |
| ***FA*** |  |  |  |  |  |
| **FAAssociationFibres** | **-0.016** | **0.004** | **-4.446** | **8.77E-06** | **2.63E-05** |
| **FAThalamicRadiations** | **-0.011** | **0.004** | **-2.861** | **0.004** | **0.006** |
| FAProjectionFibres | -0.003 | 0.004 | -0.701 | 0.484 | 0.484 |
| ***MD*** |  |  |  |  |  |
| MDAssociationFibres | 0.003 | 0.004 | 0.754 | 0.451 | 0.775 |
| MDThalamicRadiations | -0.002 | 0.003 | -0.648 | 0.517 | 0.775 |
| MDProjectionFibres | 0.001 | 0.004 | 0.200 | 0.842 | 0.842 |

***S3.1.7 DTI Individual Tract Measures***

Individual DTI tract brain measure associations with Seasonality.

p-uncorr., p-uncorrected value; p-corr., FDR p-corrected value; S.E., standard error; DF., degrees of freedom

| **Brain Imaging Measure** | **Effect Size (β)** | **S.E.** | **DF** | **t statistic** | **p-uncorr.** | **p-corr** |
| --- | --- | --- | --- | --- | --- | --- |
| ***FA*** |  |  |  |  |  |  |
| **FAintractcingulategyruspartofcingulum** | **-0.015** | **0.004** | **32800.000** | **-3.601** | **0.0003** | **0.005** |
| **FAintractinferiorfrontooccipitalfasciculus** | **-0.016** | **0.005** | **32800.000** | **-3.192** | **0.001** | **0.011** |
| **FAintractinferiorlongitudinalfasciculus** | **-0.013** | **0.005** | **32800.000** | **-2.667** | **0.008** | **0.029** |
| **FAintractposteriorthalamicradiation** | **-0.013** | **0.005** | **32800.000** | **-2.681** | **0.007** | **0.029** |
| **FAintractanteriorthalamicradiation** | **-0.012** | **0.005** | **32800.000** | **-2.495** | **0.013** | **0.038** |
| **FAintractforcepsmajor** | **-0.009** | **0.004** | **NA** | **-2.353** | **0.019** | **0.047** |
| FAintractparahippocampalpartofcingulum | -0.009 | 0.005 | 32800.000 | -1.949 | 0.051 | 0.110 |
| FAintractmediallemniscus | 0.007 | 0.005 | 32800.000 | 1.555 | 0.120 | 0.200 |
| FAintractmiddlecerebellarpeduncle | -0.006 | 0.004 | NA | -1.600 | 0.110 | 0.200 |
| FAintractforcepsminor | -0.006 | 0.004 | NA | -1.490 | 0.136 | 0.204 |
| FAintractacousticradiation | -0.003 | 0.004 | 32800.000 | -0.715 | 0.475 | 0.647 |
| FAintractsuperiorthalamicradiation | 0.003 | 0.005 | 32800.000 | 0.559 | 0.576 | 0.720 |
| FAintractuncinatefasciculus | -0.002 | 0.005 | 32800.000 | -0.452 | 0.651 | 0.752 |
| FAintractsuperiorlongitudinalfasciculus | -0.001 | 0.005 | 32800.000 | -0.291 | 0.771 | 0.826 |
| FAintractcorticospinaltract | 0.000 | 0.005 | 32800.000 | 0.069 | 0.945 | 0.945 |
| ***MD*** |  |  |  |  |  |  |
| MDintractparahippocampalpartofcingulum | 0.007 | 0.005 | 32800.000 | 1.569 | 0.117 | 0.486 |
| MDintractmediallemniscus | 0.007 | 0.004 | 32800.000 | 1.537 | 0.124 | 0.486 |
| MDintractsuperiorthalamicradiation | 0.007 | 0.004 | 32800.000 | 1.516 | 0.130 | 0.486 |
| MDintractforcepsminor | -0.007 | NA | 0.004 | -1.880 | 0.060 | 0.486 |
| MDintractcingulategyruspartofcingulum | 0.004 | 0.005 | 32800.000 | 0.875 | 0.382 | 0.573 |
| MDintractinferiorfrontooccipitalfasciculus | -0.005 | 0.005 | 32800.000 | -1.056 | 0.291 | 0.573 |
| MDintractsuperiorlongitudinalfasciculus | -0.005 | 0.005 | 32800.000 | -1.073 | 0.283 | 0.573 |
| MDintractacousticradiation | -0.005 | 0.005 | 32800.000 | -1.136 | 0.256 | 0.573 |
| MDintractposteriorthalamicradiation | -0.005 | 0.005 | 32800.000 | -1.013 | 0.311 | 0.573 |
| MDintractforcepsmajor | -0.004 | NA | 0.004 | -0.925 | 0.355 | 0.573 |
| MDintractinferiorlongitudinalfasciculus | -0.003 | 0.005 | 32800.000 | -0.659 | 0.510 | 0.690 |
| MDintractanteriorthalamicradiation | -0.003 | 0.005 | 32800.000 | -0.588 | 0.557 | 0.690 |
| MDintractmiddlecerebellarpeduncle | 0.002 | NA | 0.004 | 0.528 | 0.598 | 0.690 |
| MDintractcorticospinaltract | -0.001 | 0.005 | 32800.000 | -0.256 | 0.798 | 0.855 |
| MDintractuncinatefasciculus | -0.001 | 0.005 | 32800.000 | -0.148 | 0.882 | 0.882 |

***S3.2 Seasonality associations with brain imaging measures covarying for birth weight***

***S3.2.1 Global T1 measures***

Global T1 brain measure associations with Seasonality.

p-uncorr., p-uncorrected value; p-corr., FDR p-corrected value; S.E., standard error.

| **Brain Imaging Measure** | **Effect Size (β)** | **S.E.** | **t statistic** | **p-uncorr.** | **p-corr** |
| --- | --- | --- | --- | --- | --- |
| GlobalSurfaceArea | -0.007 | 0.004 | -1.818 | 0.069 | - |
| GlobalCorticalVolume | -0.001 | 0.004 | -0.310 | 0.757 | - |
| GlobalCorticalThickness | -0.005 | 0.004 | -1.397 | 0.162 | - |

***S3.2.2 Lobar T1 measures***

Lobar T1 brain measure associations with Seasonality.

p-uncorr., p-uncorrected value; p-corr., FDR p-corrected value; S.E., standard error.

| **Brain Imaging Measure** | **Effect Size (β)** | **S.E.** | **t statistic** | **p-uncorr.** | **p-corr** |
| --- | --- | --- | --- | --- | --- |
| ***Area*** |  |  |  |  |  |
| **FrontalArea** | **-0.010** | **0.004** | **-2.582** | **0.010** | **0.025** |
| **OccipitalArea** | **-0.011** | **0.004** | **-2.657** | **0.008** | **0.025** |
| **CingulateArea** | **-0.010** | **0.004** | **-2.422** | **0.015** | **0.026** |
| TemporalArea | -0.003 | 0.004 | -0.816 | 0.415 | 0.518 |
| ParietalArea | -0.001 | 0.004 | -0.330 | 0.741 | 0.741 |
| ***Volume*** |  |  |  |  |  |
| FrontalVolume | -0.006 | 0.004 | -1.639 | 0.101 | 0.325 |
| TemporalVolume | 0.006 | 0.004 | 1.471 | 0.141 | 0.325 |
| CingulateVolume | -0.005 | 0.004 | -1.296 | 0.195 | 0.325 |
| OccipitalVolume | -0.002 | 0.004 | -0.389 | 0.697 | 0.872 |
| ParietalVolume | 0.000 | 0.004 | -0.024 | 0.981 | 0.981 |
| ***Thickness*** |  |  |  |  |  |
| **TemporalThickness** | **0.020** | **0.005** | **4.369** | **1.25E-05** | **6.25E-05** |
| **OccipitalThickness** | **0.018** | **0.005** | **3.921** | **8.81E-05** | **0.0002** |
| **CingulateThickness** | **0.013** | **0.005** | **2.875** | **0.004** | **0.007** |
| FrontalThickness | 0.006 | 0.004 | 1.377 | 0.168 | 0.211 |
| ParietalThickness | 0.003 | 0.004 | 0.634 | 0.526 | 0.526 |

***S3.2.3 Individual T1 measures***

Individual T1 brain measure associations with Seasonality.

p-uncorr., p-uncorrected value; p-corr., FDR p-corrected value; S.E., standard error; DF., degrees of freedom

| **Brain Imaging Measure** | **Effect Size (β)** | **S.E.** | **DF** | **t statistic** | **p-uncorr.** | **p-corr** |
| --- | --- | --- | --- | --- | --- | --- |
| ***Surface area*** |  |  |  |  |  |  |
| Areaofcaudalanteriorcingulate | -0.012 | 0.004 | 21166.000 | -3.148 | 0.002 | 0.051 |
| Areaofparacentral | -0.014 | 0.006 | 21166.000 | -2.540 | 0.011 | 0.172 |
| Areaofcuneus | -0.013 | 0.006 | 21166.000 | -2.177 | 0.029 | 0.198 |
| Areaofpericalcarine | -0.013 | 0.006 | 21166.000 | -2.146 | 0.032 | 0.198 |
| Areaofsuperiorfrontal | -0.011 | 0.005 | 21166.000 | -2.228 | 0.026 | 0.198 |
| Areaofprecentral | -0.011 | 0.006 | 21166.000 | -2.048 | 0.041 | 0.209 |
| Areaofmedialorbitofrontal | -0.010 | 0.005 | 21166.000 | -1.890 | 0.059 | 0.228 |
| Areaofparahippocampal | -0.011 | 0.006 | 21166.000 | -1.890 | 0.059 | 0.228 |
| Areaoftransversetemporal | -0.007 | 0.004 | 21166.000 | -1.816 | 0.069 | 0.239 |
| Areaofposteriorcingulate | -0.009 | 0.005 | 21166.000 | -1.690 | 0.091 | 0.283 |
| Areaofparsorbitalis | -0.009 | 0.005 | 21166.000 | -1.585 | 0.113 | 0.318 |
| Areaofcaudalmiddlefrontal | -0.008 | 0.006 | 21166.000 | -1.501 | 0.133 | 0.344 |
| Areaofinsula | -0.008 | 0.005 | 21166.000 | -1.439 | 0.150 | 0.358 |
| Areaoflateraloccipital | -0.007 | 0.006 | 21166.000 | -1.341 | 0.180 | 0.376 |
| Areaofsuperiortemporal | -0.007 | 0.005 | 21166.000 | -1.334 | 0.182 | 0.376 |
| Areaoflingual | -0.007 | 0.006 | 21166.000 | -1.223 | 0.221 | 0.429 |
| Areaofparsopercularis | 0.006 | 0.005 | 21166.000 | 1.142 | 0.253 | 0.462 |
| Areaoffusiform | -0.006 | 0.005 | 21166.000 | -1.105 | 0.269 | 0.463 |
| Areaofentorhinal | 0.004 | 0.006 | 21166.000 | 0.779 | 0.436 | 0.697 |
| Areaofrostralanteriorcingulate | -0.003 | 0.004 | 21166.000 | -0.719 | 0.472 | 0.697 |
| Areaofrostralmiddlefrontal | -0.004 | 0.006 | 21166.000 | -0.745 | 0.456 | 0.697 |
| Areaofmiddletemporal | 0.004 | 0.006 | 21166.000 | 0.647 | 0.518 | 0.730 |
| Areaofinferiortemporal | 0.003 | 0.005 | 21166.000 | 0.482 | 0.630 | 0.749 |
| Areaoflateralorbitofrontal | -0.002 | 0.006 | 21166.000 | -0.450 | 0.653 | 0.749 |
| Areaofparstriangularis | -0.003 | 0.005 | 21166.000 | -0.574 | 0.566 | 0.749 |
| Areaofpostcentral | -0.003 | 0.005 | 21166.000 | -0.484 | 0.629 | 0.749 |
| Areaofsupramarginal | 0.003 | 0.005 | 21166.000 | 0.485 | 0.628 | 0.749 |
| Areaofsuperiorparietal | -0.002 | 0.006 | 21166.000 | -0.297 | 0.767 | 0.849 |
| Areaofprecuneus | 0.001 | 0.005 | 21166.000 | 0.238 | 0.812 | 0.868 |
| Areaofinferiorparietal | 0.001 | 0.005 | 21166.000 | 0.114 | 0.909 | 0.909 |
| Areaofisthmuscingulate | 0.001 | 0.005 | 21166.000 | 0.144 | 0.886 | 0.909 |
| ***Thickness*** |  |  |  |  |  |  |
| **Meanthicknessofmiddletemporal** | **0.018** | **0.005** | **21166.000** | **3.316** | **0.001** | **0.028** |
| **Meanthicknessoffusiform** | **0.018** | **0.006** | **21166.000** | **3.105** | **0.002** | **0.030** |
| **Meanthicknessofsuperiortemporal** | **0.016** | **0.006** | **21166.000** | **2.923** | **0.003** | **0.036** |
| **Meanthicknessoflingual** | **0.016** | **0.006** | **21166.000** | **2.758** | **0.006** | **0.045** |
| Meanthicknessofcuneus | 0.014 | 0.006 | 21166.000 | 2.456 | 0.014 | 0.086 |
| Meanthicknessofparahippocampal | 0.014 | 0.006 | 21166.000 | 2.397 | 0.017 | 0.086 |
| Meanthicknessofcaudalanteriorcingulate | 0.012 | 0.005 | 21166.000 | 2.322 | 0.020 | 0.090 |
| Meanthicknessofpericalcarine | 0.013 | 0.006 | 21166.000 | 2.270 | 0.023 | 0.090 |
| Meanthicknessofinferiortemporal | 0.013 | 0.006 | 21166.000 | 2.218 | 0.027 | 0.092 |
| Meanthicknessofrostralanteriorcingulate | 0.011 | 0.005 | 21166.000 | 2.082 | 0.037 | 0.116 |
| Meanthicknessoflateraloccipital | 0.012 | 0.006 | 21166.000 | 1.919 | 0.055 | 0.155 |
| Meanthicknessofmedialorbitofrontal | 0.009 | 0.006 | 21166.000 | 1.585 | 0.113 | 0.292 |
| Meanthicknessofentorhinal | 0.008 | 0.006 | 21166.000 | 1.360 | 0.174 | 0.317 |
| Meanthicknessofinsula | 0.008 | 0.006 | 21166.000 | 1.419 | 0.156 | 0.317 |
| Meanthicknessofisthmuscingulate | 0.008 | 0.006 | 21166.000 | 1.371 | 0.171 | 0.317 |
| Meanthicknessofparacentral | 0.008 | 0.006 | 21166.000 | 1.422 | 0.155 | 0.317 |
| Meanthicknessoftransversetemporal | 0.008 | 0.006 | 21166.000 | 1.413 | 0.158 | 0.317 |
| Meanthicknessofparsorbitalis | 0.005 | 0.006 | 21166.000 | 0.950 | 0.342 | 0.590 |
| Meanthicknessofinferiorparietal | 0.004 | 0.006 | 21166.000 | 0.751 | 0.453 | 0.738 |
| Meanthicknessofparstriangularis | 0.004 | 0.006 | 21166.000 | 0.713 | 0.476 | 0.738 |
| Meanthicknessofcaudalmiddlefrontal | 0.003 | 0.006 | 21166.000 | 0.569 | 0.570 | 0.771 |
| Meanthicknessoflateralorbitofrontal | 0.003 | 0.006 | 21166.000 | 0.588 | 0.557 | 0.771 |
| Meanthicknessofrostralmiddlefrontal | 0.003 | 0.006 | 21166.000 | 0.535 | 0.593 | 0.771 |
| Meanthicknessofsuperiorfrontal | 0.003 | 0.006 | 21166.000 | 0.529 | 0.597 | 0.771 |
| Meanthicknessofprecentral | 0.002 | 0.006 | 21166.000 | 0.398 | 0.691 | 0.824 |
| Meanthicknessofsupramarginal | 0.002 | 0.006 | 21166.000 | 0.398 | 0.690 | 0.824 |
| Meanthicknessofposteriorcingulate | -0.002 | 0.005 | 21166.000 | -0.309 | 0.758 | 0.855 |
| Meanthicknessofprecuneus | -0.002 | 0.006 | 21166.000 | -0.289 | 0.773 | 0.855 |
| Meanthicknessofsuperiorparietal | -0.001 | 0.006 | 21166.000 | -0.235 | 0.814 | 0.870 |
| Meanthicknessofpostcentral | -0.001 | 0.006 | 21166.000 | -0.090 | 0.928 | 0.959 |
| Meanthicknessofparsopercularis | 0.000 | 0.006 | 21166.000 | -0.007 | 0.995 | 0.995 |
| ***Volume*** |  |  |  |  |  |  |
| Volumeofcaudalanteriorcingulate | -0.006 | 0.004 | 21166.000 | -1.487 | 0.137 | 0.896 |
| Volumeofcaudalmiddlefrontal | -0.007 | 0.006 | 21166.000 | -1.196 | 0.232 | 0.896 |
| Volumeofentorhinal | 0.008 | 0.006 | 21166.000 | 1.328 | 0.184 | 0.896 |
| Volumeofinferiortemporal | 0.007 | 0.005 | 21166.000 | 1.230 | 0.219 | 0.896 |
| Volumeofisthmuscingulate | 0.006 | 0.005 | 21166.000 | 1.075 | 0.282 | 0.896 |
| Volumeofmedialorbitofrontal | -0.005 | 0.005 | 21166.000 | -0.929 | 0.353 | 0.896 |
| Volumeofmiddletemporal | 0.009 | 0.005 | 21166.000 | 1.661 | 0.097 | 0.896 |
| Volumeofparacentral | -0.007 | 0.006 | 21166.000 | -1.278 | 0.201 | 0.896 |
| Volumeofparsopercularis | 0.005 | 0.005 | 21166.000 | 0.984 | 0.325 | 0.896 |
| Volumeofpericalcarine | -0.005 | 0.006 | 21166.000 | -0.886 | 0.376 | 0.896 |
| Volumeofposteriorcingulate | -0.010 | 0.005 | 21166.000 | -1.910 | 0.056 | 0.896 |
| Volumeofprecentral | -0.006 | 0.006 | 21166.000 | -1.021 | 0.307 | 0.896 |
| Volumeofsuperiorfrontal | -0.009 | 0.005 | 21166.000 | -1.776 | 0.076 | 0.896 |
| Volumeofcuneus | -0.002 | 0.006 | 21166.000 | -0.294 | 0.768 | 0.982 |
| Volumeoffusiform | 0.001 | 0.005 | 21166.000 | 0.214 | 0.831 | 0.982 |
| Volumeofinferiorparietal | 0.002 | 0.005 | 21166.000 | 0.422 | 0.673 | 0.982 |
| Volumeofinsula | -0.002 | 0.006 | 21166.000 | -0.292 | 0.771 | 0.982 |
| Volumeoflateraloccipital | -0.002 | 0.005 | 21166.000 | -0.356 | 0.722 | 0.982 |
| Volumeoflateralorbitofrontal | 0.000 | 0.006 | 21166.000 | 0.022 | 0.982 | 0.982 |
| Volumeoflingual | 0.002 | 0.006 | 21166.000 | 0.348 | 0.728 | 0.982 |
| Volumeofparahippocampal | 0.000 | 0.006 | 21166.000 | 0.085 | 0.932 | 0.982 |
| Volumeofparsorbitalis | -0.002 | 0.005 | 21166.000 | -0.365 | 0.715 | 0.982 |
| Volumeofparstriangularis | -0.001 | 0.005 | 21166.000 | -0.118 | 0.906 | 0.982 |
| Volumeofpostcentral | -0.001 | 0.005 | 21166.000 | -0.214 | 0.830 | 0.982 |
| Volumeofprecuneus | 0.000 | 0.006 | 21166.000 | -0.048 | 0.961 | 0.982 |
| Volumeofrostralanteriorcingulate | 0.000 | 0.004 | 21166.000 | -0.085 | 0.932 | 0.982 |
| Volumeofrostralmiddlefrontal | -0.002 | 0.006 | 21166.000 | -0.354 | 0.724 | 0.982 |
| Volumeofsuperiorparietal | -0.002 | 0.006 | 21166.000 | -0.428 | 0.669 | 0.982 |
| Volumeofsuperiortemporal | 0.002 | 0.005 | 21166.000 | 0.450 | 0.652 | 0.982 |
| Volumeofsupramarginal | 0.004 | 0.005 | 21166.000 | 0.711 | 0.477 | 0.982 |
| Volumeoftransversetemporal | -0.001 | 0.005 | 21166.000 | -0.246 | 0.806 | 0.982 |

***S3.2.4 Subcortical Measures***

Subcortical T1 brain measure associations with Seasonality.

p-uncorr., p-uncorrected value; p-corr., FDR p-corrected value; S.E., standard error; DF., degrees of freedom

| **Brain Imaging Measure** | **Effect Size (β)** | **S.E.** | **DF** | **t statistic** | **p-uncorr.** | **p-corr** |
| --- | --- | --- | --- | --- | --- | --- |
| ***Volume*** |  |  |  |  |  |  |
| VolumeOfamygdala | 0.013 | 0.005 | 21166.000 | 2.422 | 0.015 | 0.108 |
| VolumeOfaccumbens | -0.005 | 0.005 | 21166.000 | -1.077 | 0.281 | 0.657 |
| VolumeOfthalamus | -0.006 | 0.006 | 21166.000 | -1.117 | 0.264 | 0.657 |
| VolumeOfcaudate | 0.003 | 0.006 | 21166.000 | 0.498 | 0.618 | 0.912 |
| VolumeOfhippocampus | 0.001 | 0.006 | 21166.000 | 0.197 | 0.844 | 0.912 |
| VolumeOfpallidum | 0.001 | 0.006 | 21166.000 | 0.133 | 0.894 | 0.912 |
| VolumeOfputamen | 0.001 | 0.006 | 21166.000 | 0.111 | 0.912 | 0.912 |

***S3.2.5 DTI Global Measures***

Global DTI tract brain measure associations with Seasonality.

p-uncorr., p-uncorrected value; p-corr., FDR p-corrected value; S.E., standard error.

| **Brain Imaging Measure** | **Effect Size (β)** | **S.E.** | **t statistic** | **p-uncorr.** | **p-corr** |
| --- | --- | --- | --- | --- | --- |
| **FATotalTracts** | **-0.017** | **0.005** | **-3.771** | **0.0002** | - |
| MDTotalTracts | -0.003 | 0.004 | -0.772 | 0.440 | - |

***S3.2.6 DTI Grouped Tract Measures***

Grouped DTI tract brain measure associations with Seasonality.

p-uncorr., p-uncorrected value; p-corr., FDR p-corrected value; S.E., standard error.

| **Brain Imaging Measure** | **Effect Size (β)** | **S.E.** | **t statistic** | **p-uncorr.** | **p-corr** |
| --- | --- | --- | --- | --- | --- |
| ***FA*** |  |  |  |  |  |
| **FAAssociationFibres** | **-0.021** | **0.005** | **-4.532** | **5.87E-06** | **1.76E-05** |
| **FAThalamicRadiations** | **-0.016** | **0.005** | **-3.426** | **0.001** | **0.001** |
| FAProjectionFibres | -0.005 | 0.005 | -1.040 | 0.298 | 0.298 |
| ***MD*** |  |  |  |  |  |
| MDProjectionFibres | 0.007 | 0.004 | 1.662 | 0.096 | 0.289 |
| MDAssociationFibres | 0.006 | 0.004 | 1.276 | 0.202 | 0.303 |
| MDThalamicRadiations | 0.001 | 0.004 | 0.301 | 0.763 | 0.763 |

***S3.2.7 DTI Individual Tract Measures***

Individual DTI tract brain measure associations with Seasonality.

p-uncorr., p-uncorrected value; p-corr., FDR p-corrected value; S.E., standard error; DF., degrees of freedom

| **Brain Imaging Measure** | **Effect Size (β)** | **S.E.** | **DF** | **t statistic** | **p-uncorr.** | **p-corr** |
| --- | --- | --- | --- | --- | --- | --- |
| ***FA*** |  |  |  |  |  |  |
| **FAintractcingulategyruspartofcingulum** | **-0.018** | **0.005** | **21166.000** | **-3.483** | 0.0005 | **0.005** |
| **FAintractinferiorfrontooccipitalfasciculus** | **-0.020** | **0.006** | **21166.000** | **-3.286** | **0.001** | **0.005** |
| **FAintractforcepsmajor** | **-0.015** | **0.005** | **0.005** | **-3.293** | **0.001** | **0.005** |
| **FAintractposteriorthalamicradiation** | **-0.019** | **0.006** | **21166.000** | **-3.125** | **0.002** | **0.007** |
| **FAintractanteriorthalamicradiation** | **-0.018** | **0.006** | **21166.000** | **-3.030** | **0.002** | **0.007** |
| **FAintractinferiorlongitudinalfasciculus** | **-0.016** | **0.006** | **21166.000** | **-2.679** | **0.007** | **0.018** |
| **FAintractforcepsminor** | **-0.011** | **0.004** | **0.005** | **-2.464** | **0.014** | **0.029** |
| FAintractuncinatefasciculus | -0.009 | 0.006 | 21166.000 | -1.474 | 0.141 | 0.238 |
| FAintractmediallemniscus | 0.008 | 0.006 | 21166.000 | 1.466 | 0.143 | 0.238 |
| FAintractparahippocampalpartofcingulum | -0.008 | 0.006 | 21166.000 | -1.320 | 0.187 | 0.280 |
| FAintractacousticradiation | -0.007 | 0.005 | 21166.000 | -1.220 | 0.222 | 0.303 |
| FAintractmiddlecerebellarpeduncle | -0.005 | NA | 0.005 | -1.038 | 0.299 | 0.374 |
| FAintractsuperiorthalamicradiation | 0.004 | 0.006 | 21166.000 | 0.625 | 0.532 | 0.613 |
| FAintractsuperiorlongitudinalfasciculus | -0.003 | 0.006 | 21166.000 | -0.527 | 0.598 | 0.641 |
| FAintractcorticospinaltract | 0.001 | 0.006 | 21166.000 | 0.182 | 0.856 | 0.856 |
| ***MD*** |  |  |  |  |  |  |
| MDintractmediallemniscus | 0.012 | 0.005 | 21166.000 | 2.292 | 0.022 | 0.329 |
| MDintractcingulategyruspartofcingulum | 0.009 | 0.006 | 21166.000 | 1.590 | 0.112 | 0.347 |
| MDintractparahippocampalpartofcingulum | 0.010 | 0.006 | 21166.000 | 1.760 | 0.078 | 0.347 |
| MDintractsuperiorthalamicradiation | 0.008 | 0.005 | 21166.000 | 1.574 | 0.116 | 0.347 |
| MDintractforcepsminor | -0.008 | NA | 0.004 | -1.903 | 0.057 | 0.347 |
| MDintractmiddlecerebellarpeduncle | 0.005 | NA | 0.005 | 1.107 | 0.268 | 0.671 |
| MDintractinferiorfrontooccipitalfasciculus | -0.004 | 0.006 | 21166.000 | -0.689 | 0.491 | 0.771 |
| MDintractsuperiorlongitudinalfasciculus | -0.005 | 0.006 | 21166.000 | -0.805 | 0.421 | 0.771 |
| MDintractuncinatefasciculus | 0.002 | 0.006 | 21166.000 | 0.397 | 0.691 | 0.771 |
| MDintractinferiorlongitudinalfasciculus | -0.003 | 0.006 | 21166.000 | -0.571 | 0.568 | 0.771 |
| MDintractcorticospinaltract | 0.002 | 0.006 | 21166.000 | 0.348 | 0.728 | 0.771 |
| MDintractacousticradiation | -0.003 | 0.006 | 21166.000 | -0.460 | 0.646 | 0.771 |
| MDintractposteriorthalamicradiation | -0.002 | 0.006 | 21166.000 | -0.435 | 0.663 | 0.771 |
| MDintractanteriorthalamicradiation | 0.002 | 0.005 | 21166.000 | 0.291 | 0.771 | 0.771 |
| MDintractforcepsmajor | 0.004 | NA | 0.005 | 0.764 | 0.445 | 0.771 |
